# Immune-Neuroendocrine Patterning and Response to Stress. A latent profile analysis in the English Longitudinal Study of Ageing

**DOI:** 10.1101/2023.07.07.23292378

**Authors:** Odessa S. Hamilton, Eleonora Iob, Olesya Ajnakina, James B. Kirkbride, Andrew Steptoe

## Abstract

Psychosocial stress exposure can disturb communication signals between the immune, nervous, and endocrine systems that are intended to maintain homeostasis. This dysregulation can provoke a negative feedback loop between each system that has high pathological risk. Here, we explore patterns of immune-neuroendocrine activity and the role of stress. Using data from the English Longitudinal Study of Ageing (ELSA), we first identified the latent structure of immune-neuroendocrine activity (indexed by high sensitivity C-reactive protein [CRP], fibrinogen [Fb], hair cortisol [cortisol], and insulin growth-factor-1 [IGF-1]), within a population-based cohort using latent profile analysis (LPA). Then, we determined whether life stress was associated with membership of different immune-neuroendocrine profiles. We followed 4,934 male and female participants with a median age of 65 years over a four-year period (2008-2012). A three-class LPA solution offered the most parsimonious fit to the underlying immune-neuroendocrine structure in the data, with 36%, 40%, and 24% of the population belonging to profiles 1 (*low-risk*), 2 (*moderate-risk*), and 3 (*high-risk*), respectively. After adjustment for genetic predisposition, sociodemographics, lifestyle, and health, higher exposure to stress was associated with a 61% greater risk of belonging to the *high-risk* profile (RRR: 1.61; 95%CI=1.23-2.12, *p*=0.001), but not the *moderate-risk* profile (RRR=1.10, 95%CI=0.89-1.35, *p*=0.401), as compared with the *low-risk* profile four years later. Our findings extend existing knowledge on psychoneuroimmunological processes, by revealing how inflammation and neuroendocrine activity cluster in a representative sample of older adults, and how stress exposure was associated with immune-neuroendocrine responses over time.

## Introduction

Communication between proinflammatory cytokines of the innate immune system with glucocorticoids and their analogs of the neuroendocrine system, is an active continuous process necessary to maintain homeostasis, even in healthy individuals.^1, 2^ Proinflammatory cytokines initiate a local inflammatory response that systemically passes through the bloodstream to endocrine and neural foci, where a number of neuroendocrine counterregulatory mechanisms are actuated, provoking a negative feedback loop.^3^ The received stimulatory signals are then transduced, leading to a complex hormonal and cytokine cascade.^4^ This integrative network between the immune, nervous, and endocrine systems is known to control physiologic processes, such as cell growth and differentiation, metabolism, and human behaviour. Dysregulation of this network has negative implications in disease aetiology,^5, 6^ with the development of a number of physical and mental ill-states, from cardiovascular disease^7^ to depression^8^, and even accelerated ageing.^9^ The high rates of chronic conditions associated with inflammatory and neuroendocrine dysregulation, along with the advancing age of the population, has provided the impetus to identify modifiable factors that could be leveraged to mitigate disease genesis; stress is one such factor.^10^

An expansive literature has elucidated the role of chronic psychosocial stress (referred to as stress hereafter) as a determinant of morbidity and mortality.^10–14^ Equally, stress has been implicated as a modulator of immune and neuroendocrine activity via psychoneuroimmunological (PNI) pathways;^15, 16^ that is, the integrative network between the nervous, endocrine, and immune systems. Therefore, if causally related to morbidity and mortality, conceivably via immune-neuroendocrine mechanisms, stress may present as a plausible preventative target to improve population health across a number of physical and mental health domains. However, the dominant position that stress disrupts immune and neuroendocrine integrity is an oversimplification of this biological pathway that fails to account for the reciprocal regulation of these transducing systems^4, 5^ and their variation among the population.^17^ Immune and neuroendocrine interactions may be intensified in the presence of stress,^15, 18^ but individuals can have highly heterogeneous patterns of immune and neuroendocrine activity, which may conflate effects and give a partial explanation for the diverse and comorbid clinical outcomes associated with stress in the literature.^10–14^

The lack of observational evidence on immune and neuroendocrine activity, as measured by their dysregulated responses, may be due to the complexity of the multidirectional exchange between these systems in response to stress.^19^ Hormonal and neuropeptide mediators that provide the link between the immune and neuroendocrine systems constitute specific axes of interactions.^3, 4, 19^ It is, thus, important to determine from a population perspective how biomarkers representing these integral systems cluster together.

The purpose of the exchange between the immune and neuroendocrine systems is to return to the physiological *status quo ante*, but many studies examine the nature of this regulation at the systemic level without considering how stress interferes with this physiological exchange.^20^ These biological responses appear to depend on stress duration and intensity, but our interest here is with chronic stress.^16, 21^ Understanding is further obfuscated by research that treats the mediators of each system as homogeneous constructs, when variation among the population is highly likely.^4^ Further, elevated inflammation and HPA-axis hyperactivity have similarities in context of stress and disease,^21^ which is paradoxical given the contrasting utility of cytokines and glucocorticoids, and the pleiotropic and redundant action between each.^1^

Owing to interindividual and intraindividual variability in biomarkers,^22^ genetic variation is another key consideration. As a major determinant of circulating immune and neuroendocrine function, genetic variation plays an important role in susceptibility to disease,^23^ and these biomarkers are of high polygenic heritability.^24^ It is, therefore, important that genetic markers are accounted for in analyses that explore immune and neuroendocrine traits.

Moreover, despite concerns of inflammaging and somatopause (i.e., age-related increases in plasma concentrations of inflammatory peptide biomarkers and the reduced expression of growth hormone secretion across age),^9^ there remains a paucity of literature on stress and immune-neuroendocrine activity in older cohorts. This demographic group is increasingly relevant from a public health perspective because of the advancing age of the population.

Classifying the different patterns of immune and neuroendocrine activity in a population-based cohort of older adults, quantifying their prevalence, and identifying which profiles are most strongly associated with long-term stress exposure could be beneficial for three reasons. First, it may help to elucidate some of the present uncertainty about immune and neuroendocrine patterning. Second, it could contribute to more targeted preventative treatments and novel therapeutic strategies, such as the identification of biomarkers that characterise patients into subgroups most likely to benefit from cytokine-mediated pharmacological treatments, or the design of more personalised clinical trials through targeted recruitment. Third, it could be a resource for the formulation of more robust hypotheses for future research exploring stress models in immune and neuroendocrine activity, and their subsequent roles in human health and behaviour.

We sought to address these issues in a UK cohort of community-dwelling older adults, to classify and quantify distinct immune and neuroendocrine profiles, and to determine the longitudinal association between psychosocial stress and the revealed profiles. To represent these interrelated, molecular pathways, we selected two acute-phase reactants (i.e., C-reactive protein and fibrinogen) and two hormones; one catabolic (i.e., hair cortisol), the other anabolic (i.e., insulin-like growth factor-1). We expected heterogeneous patterns of immune and neuroendocrine activity, with two to three subgroups emerging from the data. We also expected psychosocial stress to be longitudinally associated with more adverse immune and neuroendocrine patterns four years later.

## Method

### Study Design

This prospective cohort study used fully anonymised data from the English Longitudinal Study of Ageing (ELSA), a nationally representative, multidisciplinary prospective observational study of the English population aged 50 years and older.^25^ The present study used data from ELSA participants at wave 4 (2008), who were followed up four years later at wave 6 (2012).

### Exposures

Psychosocial stress was assessed at wave 4 (2008), measured as a composite score on a scale from no stressful life events to the experience of six stressors. Thus, we estimated an ordinal score as the summation of the presence of six binary stressors. We dichotomised this score (low versus high) at the median. Despite this median split, there is an unequal distribution of participants in each group due to the limited number of integer values of this score (0-6):

1. **Financial Strain.** Binary:- the perceived chance of not having enough financial resources in the future to meet needs; categorised by 0; 1-39; 40-60; 61-99; 100% and dichotomised at >60%.
2. **Care Giving.** Binary:- being an informal caregiver to an adult who is sick or frail, in the past week, or during the last month while being in receipt of Carer’s Allowance.
3. **Disability.** Binary:- has one or more difficulties mobilising (i.e., walking 100 yards; sitting 2-hours; rising from chairs after sitting long periods; climbing stairs; stooping, kneeling, crouching; reaching or extending arms above shoulders; pulling or pushing large objects; lifting or carrying objects over 10 pounds; picking-up a 5p coin).
4. **Illness.** Binary:- has a longstanding illness or health condition that limits activity.
5. **Bereavement.** Binary:- experienced the death of a parent, spouse, or partner within the past two years.
6. **Divorce.** Binary:- experienced divorce or the breakdown of a long-term relationship within the past two years.

### Outcomes

Immune and neuroendocrine biomarkers measured at wave 6 (2012) included high-sensitivity plasma C-reactive protein (CRP; mg/L), plasma fibrinogen (Fb; g/L), serum insulin-like growth factor-1 (IGF-1; mmol/L) and hair cortisol (cortisol; pg/mg). The complete immunoassay procedure can be found in Supplementary Materials (SM) 1. Blood samples deemed insufficient or unsuitable (e.g., haemolysed; received >5 days post-collection) were discarded. Exclusion criteria included coagulation, haematological disorders, being on anticoagulant medication or having a history of convulsions (SM 1). We then conducted a latent profile analysis (LPA) on these immune and neuroendocrine biomarkers (see below).

### Covariates (Wave 4)

Factors likely to confound analyses were selected *a priori* (see Figure S1 for the Directed Acyclic Graph), including *demographic variables*: age (≥50 years); sex (male; female); *socioeconomic variables*: education (categorised into higher education; primary/secondary/tertiary education; or alternative/none); occupational social class (a three-category version of the National Statistics Socio-Economic Classification:^26^ managerial and professional; intermediate; routine and manual); *lifestyle variables*: smoking status (binary:- non-smokers/ex-smokers or smokers); alcohol consumption (binary:- low <3 or high ≥3 day weekly); physical activity (binary:- sedentary or moderate/vigorous weekly activity); *genetic variables*: polygenic scores (PGS) for CRP, cortisol, and IGF-1 (methods later described) and 10 principal components to account for population stratification; *biomarkers*: baseline (wave 4) CRP, fibrinogen, and IGF-1 entered into the LPA; (Figures S2-3); *binary health indicator*: any self-reported physician diagnosis of chronic lung disease, coronary heart disease, abnormal heart rhythm, heart murmur, congestive heart failure, angina, hypertension, diabetes, cancer, Parkinson’s, Alzheimer’s, dementia, asthma, arthritis, osteoporosis, psychiatric disorder.

### Genetic data

Using PLINK and PRSice software, PGS for CRP, cortisol, and IGF-1 were calculated using summary statistics from genome-wide association studies (GWAS) from the UK Biobank.^27^ A single *p*-value threshold of 0.001 was used for all PGSs to limit multiple testing, while maximising their potential predictive ability (further detail can be found in SM2).

#### Imputation

Missingness ranged from 0.00-52.26%, with cortisol having the greatest proportion of missingness, and other variables having less than 37% missing (Table S1). Given the possibility of bias in the complete case analyses,^28^ missing values on exposures, covariates, and outcomes were imputed using missForest.^29^ This is an algorithm based on Random Forests, a machine learning iterative imputation method in R v.4.2.0: RStudio v.2022.02.2. We did not impute missing genetic data; participants without genetic information were excluded from the analyses, as detailed in the analytic sample formation (Figure S4). The imputation of the missing values yielded minimal error for continuous variables (Normalized Root Mean Squared Error=0.02%) and categorical variables (proportion of falsely classified=0.07%). Imputed and observed data were comparable in terms of their summary distributions on participant characteristics (Table S1).

### Statistical Analyses

First, we reported baseline (wave 4) characteristics, expressed as means and proportions. Fibrinogen was normally distributed but logarithmic transformation was performed on CRP, Cortisol, and IGF-1 values because of their originally skewed distribution.

Second, we conducted an LPA to determine patterns of immune and neuroendocrine activity at both waves. The optimal number of profiles was identified using a stepwise approach. Starting with a single-profile model, additional profiles were added to determine whether it improved the model fit (further detail can be found in SM3). Once the number of latent profiles was determined, each individual in the sample was then assigned to a cluster for which they had the largest posterior probability (i.e., the profile they most likely belonged to).

Third, we used multinomial logistic regression to investigate the association between psychosocial stress at wave 4 (2008) and the probability of immune and neuroendocrine profile membership at wave 6 (2012). Results were presented as relative risk ratios (RRR), with standard errors (SE) and 95% confidence intervals (95% CI). Analyses were two-tailed. Models with different sets of covariates were fitted to understand their role in the association between stress and immune and neuroendocrine profiles. Model 1 was unadjusted. Model 2 adjusted for baseline immune and neuroendocrine profiles. Model 3 additionally adjusted for *demographic* and *genetic* variables. Model 4 adjusted for all covariates. All data analyses were conducted in Stata 17.1 (StataCorp, TX, USA).

### Sensitivity Analyses

We conducted six sensitivity analyses to examine the robustness of our findings. First, to ensure associations were not dependent on the binary classification of stress, analyses were repeated using an ordinal score of stress (reported as unstandardized regression coefficients with standard errors [SE]). Second, to reveal any differences in stress exposure on profile membership, regressions were repeated using each of the six psychosocial stressors independently. Third, individuals who were disabled or with longstanding limiting illness were more likely to be immunosuppressed given anti-inflammatory prescriptions, thus altering immune and neuroendocrine activity. Therefore, we reconstructed our stress index excluding these measures, then reran our analyses to quantify the extent to which they could have biased our results. Fourth, due to the potentially confounding effects of inflammaging and somatopause,^9^ along with known differences in stress associations across age,^30^ the moderating effect of age was tested (dichotomised by mean age [≥65 years]). Fifth, because of known sex differences in biomarker activity,^31^ effect modification by sex was tested. Finally, we compared results from our imputed analyses with a complete case analysis (CCA) to understand the potential impact of different approaches to deal with missing data on the results. The analytical sample formation for CCA is illustrated in Figure S5.

## Results

Of the 6,512 core respondents, 1,578 had missing genetic data (required to control for potential genetic confounding, see below), leaving a final analytic sample of 4,934 (Figure S4). Participant characteristics of the analytic sample were materially unchanged from participants in the core sample (Table S1) and are shown in Table 1. CRP was linearly correlated with fibrinogen (*r*=0.706); cortisol (*r*=0.273); and IGF-1 (*r*=-0.163), as fibrinogen was with cortisol (*r*=0.176; all at *p*<0.001; Table S2). Participants, male (∼45%) and female (∼55%), with a median age of 65 years old (interquartile range: 59-72; M_age_=66.31; ±9.35; _range_50-99) were followed over a four year period (2008-2012). Most were non-smokers (87.27%) and consumed alcohol less than three days a week (64.27%), and almost two thirds were sedentary (72.88%). There was a fairly equal educational (Higher - 32.12%; Primary/Secondary/Tertiary - 31.29%; Alternative/None - 36.58%) and occupational social class divide (Managerial/Professional - 36.28%; Intermediate Occupations - 25.62%; Routine/Manual - 38.10%). There were 8,083 unique documented stress experiences (Figure S6). Approximately 13% of the sample experienced a high level of stress, and this high stress group tended to be younger, female, smokers, who drank less than three alcoholic drinks a week (Table 1). As it pertains to each independent stressor, 17.02% of the sample experienced financial strain, 7.01% were informal carers, 45.80% had difficulty mobilising, 31.46% had a limiting longstanding illness, 40.86% were bereaved, and 9.18% were divorcees (Figure S7).

**Table 1.**
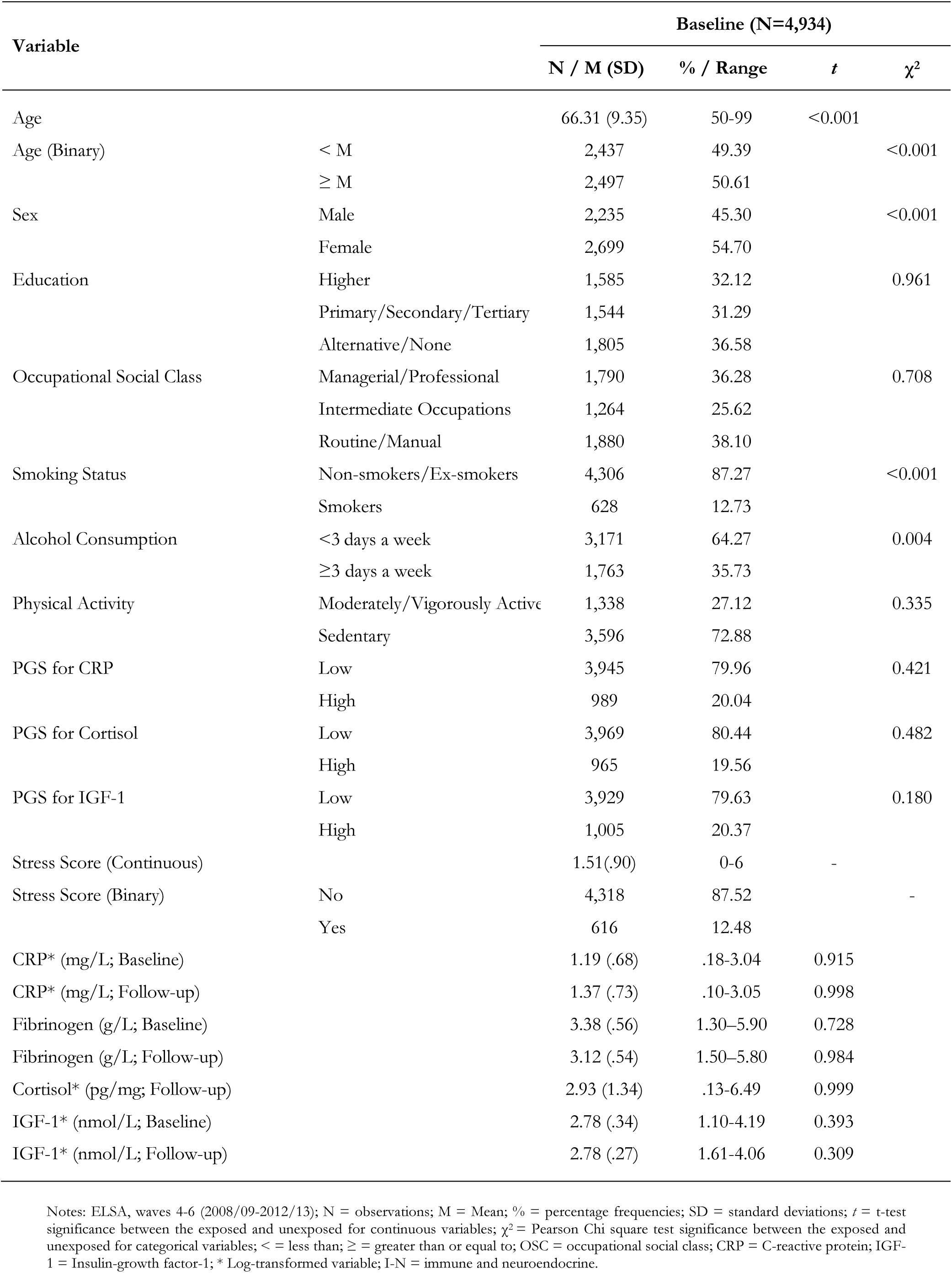
Sample characteristics

| Variable |  | Baseline (N=4,934) |  |  |  |
| --- | --- | --- | --- | --- | --- |
| | | N / M (SD) | % / Range | *t* | $\chi^2$ |
| Age |  | 66.31 (9.35) | 50-99 | <0.001 |  |
| Age (Binary) | < M | 2,437 | 49.39 |  | <0.001 |
|  | ≥ M | 2,497 | 50.61 |  |  |
| Sex | Male | 2,235 | 45.30 |  | <0.001 |
|  | Female | 2,699 | 54.70 |  |  |
| Education | Higher | 1,585 | 32.12 |  | 0.961 |
|  | Primary/Secondary/Tertiary | 1,544 | 31.29 |  |  |
|  | Alternative/None | 1,805 | 36.58 |  |  |
| Occupational Social Class | Managerial/Professional | 1,790 | 36.28 |  | 0.708 |
|  | Intermediate Occupations | 1,264 | 25.62 |  |  |
|  | Routine/Manual | 1,880 | 38.10 |  |  |
| Smoking Status | Non-smokers/Ex-smokers | 4,306 | 87.27 |  | <0.001 |
|  | Smokers | 628 | 12.73 |  |  |
| Alcohol Consumption | <3 days a week | 3,171 | 64.27 |  | 0.004 |
|  | ≥3 days a week | 1,763 | 35.73 |  |  |
| Physical Activity | Moderately/Vigorously Active | 1,338 | 27.12 |  | 0.335 |
|  | Sedentary | 3,596 | 72.88 |  |  |
| PGS for CRP | Low | 3,945 | 79.96 |  | 0.421 |
|  | High | 989 | 20.04 |  |  |
| PGS for Cortisol | Low | 3,969 | 80.44 |  | 0.482 |
|  | High | 965 | 19.56 |  |  |
| PGS for IGF-1 | Low | 3,929 | 79.63 |  | 0.180 |
|  | High | 1,005 | 20.37 |  |  |
| Stress Score (Continuous) |  | 1.51(.90) | 0-6 | - |  |
| Stress Score (Binary) | No | 4,318 | 87.52 |  | - |
|  | Yes | 616 | 12.48 |  |  |
| CRP* (mg/L; Baseline) |  | 1.19 (.68) | .18-3.04 | 0.915 |  |
| CRP* (mg/L; Follow-up) |  | 1.37 (.73) | .10-3.05 | 0.998 |  |
| Fibrinogen (g/L; Baseline) |  | 3.38 (.56) | 1.30–5.90 | 0.728 |  |
| Fibrinogen (g/L; Follow-up) |  | 3.12 (.54) | 1.50–5.80 | 0.984 |  |
| Cortisol* (pg/mg; Follow-up) |  | 2.93 (1.34) | .13-6.49 | 0.999 |  |
| IGF-1* (nmol/L; Baseline) |  | 2.78 (.34) | 1.10-4.19 | 0.393 |  |
| IGF-1* (nmol/L; Follow-up) |  | 2.78 (.27) | 1.61-4.06 | 0.309 |  |

Notes: ELSA, waves 4-6 (2008/09-2012/13); N = observations; M = Mean; % = percentage frequencies; SD = standard deviations; *t* = t-test significance between the exposed and unexposed for continuous variables; $\chi^2$ = Pearson Chi square test significance between the exposed and unexposed for categorical variables; < = less than; ≥ = greater than or equal to; OSC = occupational social class; CRP = C-reactive protein; IGF-1 = Insulin-growth factor-1; \* Log-transformed variable; I-N = immune and neuroendocrine.

### Latent Profile Analysis of Immune and Neuroendocrine Biomarkers

A three-profile model of immune and neuroendocrine biomarkers provided the most parsimonious fit to biomarker data at wave 6 (Table S3; Figures S8 [a-g]), after which there were limited returns in AIC and BIC value (Figure S9); entropy was above 0.80 (Figure S10); the mean posterior probabilities did not exceed 0.70; each profile comprised more than 5% of participants (Figure S11; Table S3); and each profile was theoretically meaningful. The most common profile was 2 (40%), followed by profile 1 (36%), then profile 3 (24%; Figure S12). Profile 1 (M_age_=64.16; ±7.77; 36% of the sample) was defined as ‘*low-risk*’ as it was characterised by those having low CRP, low fibrinogen, low cortisol, and high IGF-1. Profile 2 (M_age_=66.59; ±9.38; 40% of the sample) was the modal group, and consisted of individuals with moderate CRP, fibrinogen, cortisol, and IGF-1 levels, which was defined as ‘*moderate-risk*’. Finally, profile 3 (M_age_=69.03; ±10.62; 24% of the sample) was marked by a high probability of high CRP, high fibrinogen, high cortisol, and low IGF-1, so this group was defined as ‘*high-risk*’ (Figure 1).

**Figure 1.**
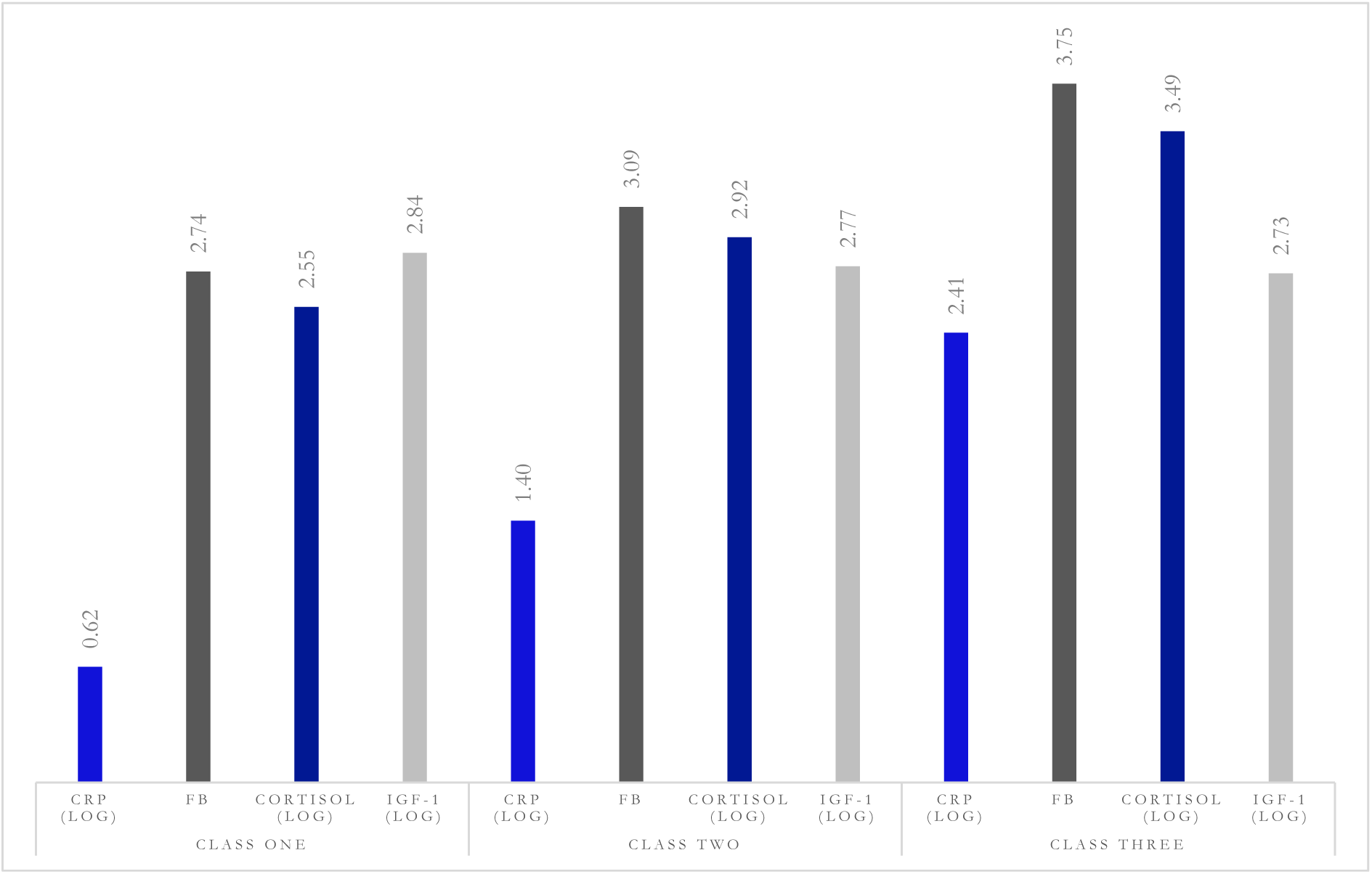
The mean levels of immune and neuroendocrine biomarkers for a three-profile solution

### Stress and Profile Membership of Immune and Neuroendocrine Biomarkers

In the unadjusted model, greater stress was associated with the probability of being in the *high-risk* profile versus *low-risk* profile (Model [M] 1: RRR=1.34, 95%CI=1.08-1.66, *p*=0.008). This persisted after adjustment for baseline immune and neuroendocrine profiles (M2: RRR=1.42, 95%CI=1.10-1.83, *p*=0.007), further adjustment for *demographic* and *genetic variables* (M3: RRR=1.80, 95%CI=1.39-2.35, *p*<0.001), and in our fully adjusted model, the risk of a *high-*immune and neuroendocrine profile (was *low-risk* profile) was 1.6 times higher in the group exposed to high levels of stress compared with participants with lower stress exposure (M4: RRR=1.61, 95%CI=1.23-2.12, *p*=0.001). In the fully adjusted model, however, stress was not associated with the probability of being in the *moderate-risk* profile versus *low-risk* profile (Model M4: RRR=1.10, 95%CI=0.89-1.35, *p*=0.401; Table 2). To understand the role of specific confounding factors with greater nuance, results with incremental model adjustment can be found in the supplement (Table S4). There was evidence of negative confounding by *demographic* and *genetic variables,* which increased the RRR by 38% (M3: RRR=1.80, 95%CI=1.39-2.35, *p*<0.001), and by *health* variables, which increased the RRR by 20% (M3c: RRR=1.81, 95%CI=1.39-2.36, *p*<0.001).

**Table 2.**
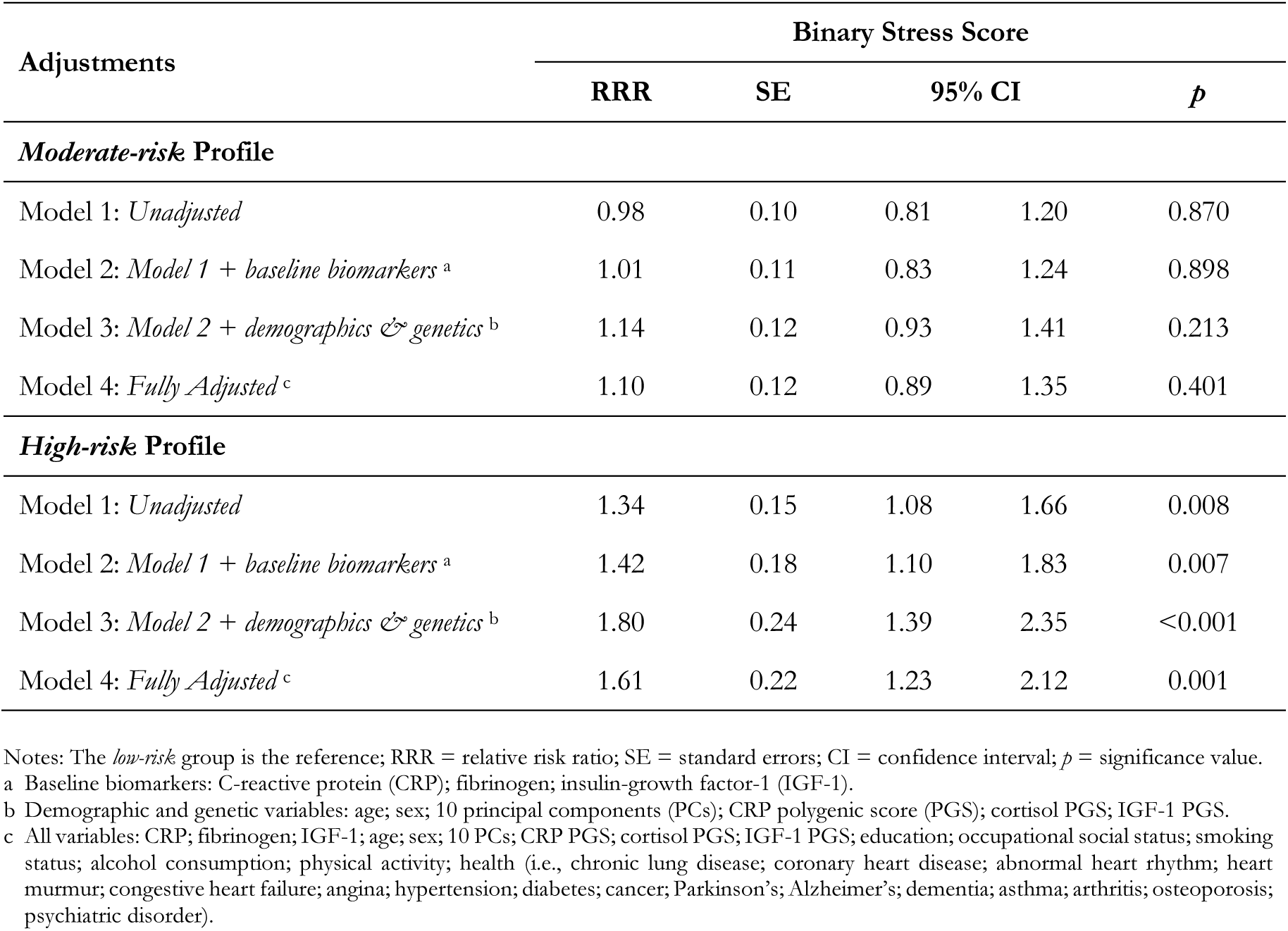
Longitudinal associations of stress with immune and neuroendocrine biomarker profiles (N=4,934)

### Sensitivity Analyses

First, results were consistent when we used a continuous classification of psychosocial stress. For each single increase in the stress score, individuals were 19% more likely to be in the *high-risk* immune and neuroendocrine profile versus the *low-risk* profile in our fully adjusted model (M4: RRR=1.19, 95%CI=1.23-2.12, *p*=0.001; Table S5). Second, when individual stressors were tested against immune and neuroendocrine profile membership, we found that financial strain (M4: RRR=1.59, 95%CI=1.25-2.01, *p*<0.001), limiting longstanding illness (M4: RRR=1.34, 95%CI=1.10-1.65, *p*=0.005), and bereavement (M4: RRR=1.26, 95%CI=1.04-1.52, *p*=0.016) were each associated with belonging to the *high-risk* profile, as compared with the *low-risk* profile in fully adjusted models. Financial strain and bereavement showed gradients in risk, as each were associated with *high-* and *moderate-risk* profile membership. Caregiving and divorce were not associated with differences in profile membership, while disability was associated with a 30% lower risk of belonging to the high-risk profile (Tables S6[a-f]). Third, the stress index that excluded both disability and limiting long standing illness had higher relative risk coefficients than the primary composite score (M4: RRR=1.71, 95%CI=1.32-2.22, *p*<0.001), consistent with the previous observation with respect to disability (Table S7). Fourth, we found no evidence of differences in the association between stress and biomarker profile membership between younger and older age groups (interaction *p=*0.913), although relative risk coefficients were substantially larger for those aged 65 and older (Table S8[a-b]). Fifth, similar to age, there was no interaction (*p=*0.239) nor difference in the risk profile between the sexes when results were stratified by sex (Tables S9[a-b]). Finally, we observed similar mean levels of immune and neuroendocrine biomarkers for a three-profile solution in a CCA (Figures S13-14) as compared with the main imputed data (Figure S8c). Re-analysis of the association between stress and profile membership in the CCA sample yielded similar results (Table S10).

## Discussion

In a large nationally representative sample of UK older adults, we used multiple biomarkers in a latent profile analysis to provide a comprehensive characterisation of physiological activity across the integrative network of the immune, nervous, and endocrine systems. We found longitudinal evidence of an overall association between stress and the risk of high versus low immune and neuroendocrine profile membership four years later. Associations remained significant after accounting for polygenic markers of immune and neuroendocrine activity, and a range of demographic, socioeconomic, lifestyle, and health factors. There was, however, no consistent gradient in risk as there was no significant difference in stress levels between *low-* and *moderate-risk* profiles, nor were there differences in the association between stress and immune-neuroendocrine profile activity by age or sex. Stress associated with financial strain was the strongest independent determinant of belonging to the *high-risk* immune and neuroendocrine profile, followed by limiting longstanding illness and bereavement. Furthermore, financial strain and bereavement showed gradients in risk. In contrast, disability was associated with a lower risk for *moderate*- and *high-risk* profile membership (vs *low-risk*).

As noted elsewhere,^32^ the biological responses to stress exposure are multiphasic, where we see the stimulation or suppression of immune and neuroendocrine activity, or both simultaneously,^33^ with the direction of effect depending on the biomarker being evaluated.^3, 34^ We addressed the complexity of immune and neuroendocrine interconnectivity by using latent profile analyses to identify distinct typologies of activity. Variability was revealed within the derived profiles, and highlights why the evaluation of single biomarkers can obfuscate understanding of stress exposure. Though each biomarker has a unique role in maintaining health, functionally they are involved in proliferation, differentiation, migration, and apoptosis of targeted cells.^35^ They are characterised by interrelated pleiotropic, synergistic, and redundant actions that have afferent and efferent functional components.^6^ When this dynamic process is dysregulated, it leads to varying concentrations of circulating biomarkers^4^ that can contribute to diversity in disease sequelae.^15, 18, 36^ This can make prediction more challenging and the interpretation of single biomarkers less intuitive, particularly because issues of multicollinearity mean that biomarkers are best modelled independently in regressions.^34^ Our latent variable modelling approach, similar to an earlier study of American adults,^37^ allowed for a synchronised assessment of a diverse set of biomarkers. However, these studies are not comparable because the selection of immune and neuroendocrine biomarkers differed. Even so, our derived profiles lend support to earlier experimental research that indicate symmetry between biomarkers of the immune, nervous, and endocrine systems,^2, 3, 19, 38^ and our results confirm that that biomarkers are, on average, temporally stable, despite individual trajectories varying widely.^34^

The incremental rise in mean fibrinogen and cortisol levels from profile one to three, aligns with increases in mean CRP, which is consistent with earlier evidence on the synchronised physiological exchange between their respective systems to maintain homeostasis.^1^ However, the unexpected moderate decline in IGF-1 between each of the derived profiles is notable. The reasons for this is unclear given the well documented covariance between each represented system in the LPA.^2, 3, 34, 39^ As part of a coordinated systemic regulatory mechanism that facilitates a dynamic cellular microenvironment, proinflammatory cytokines can induce a state of resistance in hormonal secretion, including in IGF-1.^3^ This can attenuate the mitogenic effect of IGF-1, but can also have anti-proliferative effects on IGF-1,^2^ which should be reflected here. The reason for the blunted effect of IGF-1 seen in the present study, is conceivably because IGF-1 secretion is sensitive to nutritional and endocrine control, such that hormonal resistance is rendered maladaptive by pharmacologic use and dietary choices;^40^ neither of which were measured here. In addition, O’Connor and colleagues (2008)^2^ suggest that cellular responses can vary tremendously depending on ligand origin and concentration, the number of cell receptors, and signalling kinetics post receptor activation, not to mention extracellular control of IGF-1, which is a second mode of regulation.

It is also clear from converging lines of evidence that different stressors have different predictive power.^14, 15, 17, 21^ There was some evidence to support this in the present study, with the largest effect sizes observed following financial stress, but given the overlap of CI, there is not strong associative differentiation. Even so, Hamilton and Steptoe’s (2022)^34^ recent observational study revealed idiosyncrasies in the role of different socioeconomic stressors in CRP, fibrinogen, IGF-1, and white blood cell count (WBCC/leukocytes). Part of the challenge is in establishing a ‘*hierarchy of stress*’ to determine which psychosocial stressors are most problematic; distinguishing between rare acute stressors that have high clinical risk and everyday stressors that create chronic risk and contribute more to overall disease burden in the population. The present study takes a step toward this purpose, and while we used an LPA to look at immune and neuroendocrine patterning here, future study would benefit from a more comprehensive stress score that is also submitted to LPA to see how stress clusters in the population.

Our results extend previous evidence on psychoneuroimmunological processes,^15, 17, 21^ by showing that stress exposure is associated with a greater probability of *high-risk* immune and neuroendocrine profile membership, irrespective of genetic propensity. This is an important feature of our study, and a methodological advance over previous research given that genetic factors can affect the magnitude of the immune and neuroendocrine response.^24^ Inter-individual variability in biomarker concentrations and their respective binding proteins are partly the result of polymorphic variations in respective genes, while genes encoding biomarkers are candidate loci for diseases with an inflammatory basis.^24^ Moreover, CRP,^41^ fibrinogen,^41^ cortisol,^42^ and IGF-1^43^ each have high heritability, which can be understood as the proportion of the total variation of the trait that can be attributed to unobserved genetic effects.^44^ Therefore, while single nucleotide polymorphisms (SNPs) associated with each biomarker only explained a small proportion of the variance in our phenotypic associations, it is plausible that they confounded earlier evidence, such that their omission inflated effect sizes.

Our study has several strengths. To our knowledge this is the first study to explore how stress is related to immune and neuroendocrine profile membership. The application of a latent profile approach and the prospective nature of the study facilitated an exploration into the temporal direction of stress associations with population-level configurations of immune and neuroendocrine biomarker activity with increased specificity. LPA was chosen over other traditional clustering methods because it identifies subgroups of individuals with similar biomarker activity.^45^ Dichotomising the ordinal stress score reduced the influence of its non-normality, quasi-continuous quality, and limited the chance of underestimated correlations and an inflation of Type II errors (i.e., false negatives). Therefore, it offered more meaningful results, despite the potential loss of power. Another key strength is in our use of a well-powered, well-characterised cohort that offers precise estimates of objective, systematically measured, interrelated biomarkers.^25^ In the presence of nonlinearity and interactions missForest outperforms prominent imputation methods, such as multivariate imputation by chained equations and k-nearest neighbours in all metrics.^29^

We do, however, note some important caveats. We cannot claim causality. Given the observational nature of the study, our results might be subject to residual confounding or over-adjustment. While the fibrinogen PGS was not available, a strong genetic correlation with CRP has been documented elsewhere,^41^ and PGS for CRP was accounted for in analyses. Similarly, baseline cortisol was unavailable, although follow-up cortisol was correlated with CRP and fibrinogen (Table S2); both adjusted for at baseline. The self-reported nature of the stress score may have introduced some measurement error to the results, and there is an assumption in the stress measure that different exposures carry equal weight but this is typically not so. Given that ELSA participants are 99% White, and ethnic groups are said to experience higher levels of stress,^46^ their absence in the present study is a considerable limitation. Crucially, immune and neuroendocrine activation involves a constellation of cells that interact and create a microenvironment that promotes disease, but here we include a relatively small number of biomarkers to represent this complex network.

## Conclusion

The synergistic immune and neuroendocrine response to stress represents an important target for clinical intervention. Intervening on these processes could alter the course of disease.^47^ We examined multivariate biomarkers, including CRP, fibrinogen, cortisol, and IGF-1, using empirically derived data reduction techniques to uncover subgroup differences in how immune and neuroendocrine biomarkers pattern together. It proved an effective method to explore the complex series of reactions across the immune, nervous, and endocrine systems. Because stress was positively associated with the derived immune and neuroendocrine profiles, our results support that exposure to high levels of stress can actuate a cascade of complex central and peripheral physiological events that has previously been linked to pathology, sub-clinical illness, and debility.

## Data Availability

All data produced in the present work are contained in the manuscript.

## Acknowledgement and Funding

**Funding.** ELSA is funded by the National Institute on Aging (R01AG017644), and by UK Government Departments coordinated by the National Institute for Health and Care Research (NIHR). The data are linked with the UK Data Archive and freely available through the UK data services and can be accessed here: <u>discover.ukdataservice.ac.uk</u>. AS is the director of the study. OSH is supported by the Economic and Social Research Council (ESRC), and the Biotechnology and Biological Sciences Research Council (BBSRC), UCL Soc-B Doctoral Studentship (ES/P000347/1). **Data Sharing.** The data are deposited in the UK Data Archive and freely available through the UK Data Service (SN 8688 and 5050) and can be accessed here: <u>discover.ukdataservice.ac.uk</u>. **Ethical Approval.** The National Research Ethics Service (London Multicentre Research Ethics Committee [MREC/01/2/91] <u>nres.npsa.nhs.uk</u>) granted ethical approval for each of the ELSA waves. All participants provided informed consent, and research was performed in accordance with research and data protection guidelines. **Contributorship.** Study funding was secured by AS. Conception and planning by OSH and AS. OSH and EI designed the statistical analysis plan. OSH and OA and prepared the data. OSH performed data analyses, then interpreted results with input from all authors. All authors had access to the data, take responsibility for data integrity, the accuracy of analysis, and its interpretation. All authors act as guarantors, and critically appraised the manuscript drafted by OSH for submission.

## References

1. Shimba, A., Ejima, A. & Ikuta, K. Pleiotropic Effects of Glucocorticoids on the Immune System in Circadian Rhythm and Stress. Frontiers in Immunology 12, (2021).

2. O’Connor, J. C. et al. Regulation of IGF-I function by proinflammatory cytokines: At the interface of immunology and endocrinology. Cellular Immunology 252, 91–110 (2008).

3. Taub, D. D. Neuroendocrine Interactions in the Immune System. Cell Immunol 252, 1–6 (2008).

4. Chikanza, I. C. & Grossman, A. B. Reciprocal Interactions Between the Neuroendocrine and Immune Systems During Inflammation. Rheumatic Disease Clinics of North America 26, 693–711 (2000).

5. Dantzer, R. Neuroimmune Interactions: From the Brain to the Immune System and Vice Versa. Physiological Reviews 98, 477–504 (2017).

6. Kany, S., Vollrath, J. T. & Relja, B. Cytokines in Inflammatory Disease. Int J Mol Sci 20, 6008 (2019).

7. Iob, E. & Steptoe, A. Cardiovascular Disease and Hair Cortisol: a Novel Biomarker of Chronic Stress. Curr Cardiol Rep 21, 116 (2019).

8. Iob, E., Kirschbaum, C. & Steptoe, A. Persistent depressive symptoms, HPA-axis hyperactivity, and inflammation: the role of cognitive-affective and somatic symptoms. Molecular Psychiatry 1–11 (2019) doi:10.1038/s41380-019-0501-6.

9. Wagner, K.-H., Cameron-Smith, D., Wessner, B. & Franzke, B. Biomarkers of Aging: From Function to Molecular Biology. Nutrients 8, 338 (2016).

10. Acabchuk, R. L., Kamath, J., Salamone, J. D. & Johnson, B. T. Stress and chronic illness: The inflammatory pathway. Social Science & Medicine 185, 166–170 (2017).

11. Steptoe, A. & Kivimäki, M. Stress and cardiovascular disease. Nature Reviews Cardiology 9, 360–370 (2012).

12. Batty, G. D. et al. Psychosocial factors and hospitalisations for COVID-19: Prospective cohort study based on a community sample. Brain, Behavior, and Immunity 89, 569–578 (2020).

13. Hamer, M., Kivimaki, M., Stamatakis, E. & Batty, G. D. Psychological distress and infectious disease mortality in the general population. Brain, Behavior, and Immunity 76, 280–283 (2019).

14. Cohen, S., Janicki-Deverts, D. & Miller, G. E. Psychological Stress and Disease. JAMA 298, 1685–1687 (2007).

15. Kiecolt-Glaser, J. K., McGuire, L., Robles, T. F. & Glaser, R. Psychoneuroimmunology: Psychological influences on immune function and health. Journal of Consulting and Clinical Psychology 70, 537–547 (2002).

16. Johnson, J. D., Barnard, D. F., Kulp, A. C. & Mehta, D. M. Neuroendocrine Regulation of Brain Cytokines After Psychological Stress. Journal of the Endocrine Society 3, 1302–1320 (2019).

17. Steptoe, A., Hamer, M. & Chida, Y. The effects of acute psychological stress on circulating inflammatory factors in humans: A review and meta-analysis. *Brain*, Behavior, and Immunity 21, 901–912 (2007).

18. Hamilton, O. S., Cadar, D. & Steptoe, A. Systemic inflammation and emotional responses during the COVID-19 pandemic. Transl Psychiatry 11, 1–7 (2021).

19. Ménard, C., Pfau, M. L., Hodes, G. E. & Russo, S. J. Immune and Neuroendocrine Mechanisms of Stress Vulnerability and Resilience. Neuropsychopharmacol 42, 62–80 (2017).

20. Andreassen, M., Frystyk, J., Faber, J. & Kristensen, L. O. GH activity and markers of inflammation: a crossover study in healthy volunteers treated with GH and a GH receptor antagonist. European Journal of Endocrinology 166, 811–819 (2012).

21. Segerstrom, S. C. & Miller, G. E. Psychological Stress and the Human Immune System: A Meta-Analytic Study of 30 Years of Inquiry. Psychological Bulletin 130, 601–630 (2004).

22. de Maat, M. P. M. et al. Interindividual and Intraindividual Variability in Plasma Fibrinogen, TPA Antigen, PAI Activity, and CRP in Healthy, Young Volunteers and Patients With Angina Pectoris. *Arteriosclerosis*, Thrombosis, and Vascular Biology 16, 1156–1162 (1996).

23. Frank, P., Ajnakina, O., Steptoe, A. & Cadar, D. Genetic susceptibility, inflammation and specific types of depressive symptoms: evidence from the English Longitudinal Study of Ageing. Translational Psychiatry 10, 1–9 (2020).

24. Prins, B. P. et al. Genome-wide analysis of health-related biomarkers in the UK Household Longitudinal Study reveals novel associations. Sci Rep 7, 11008 (2017).

25. Steptoe, A., Breeze, E., Banks, J. & Nazroo, J. Cohort Profile: The English Longitudinal Study of Ageing. Int J Epidemiol 42, 1640–1648 (2013).

26. ONS, O. for N. S. The National Statistics Socio-economic classification (NS-SEC) - Office for National Statistics. https://www.ons.gov.uk/methodology/classificationsandstandards/otherclassifications/thenationalstatisticssocioeconomicclassificationnssecrebasedonsoc2010 (2010).

27. Ajnakina, O. & Steptoe, A. THE ENGLISH LONGITUDINAL STUDY OF AGEING (ELSA) POLYGENIC SCORES 2019. https://www.ucl.ac.uk/epidemiology-health-care/sites/epidemiology_health_care/files/elsa_gwas_pgs-apr2019.pdf (2019).

28. Sterne, J. A. C. et al. Multiple imputation for missing data in epidemiological and clinical research: potential and pitfalls. BMJ 338, b2393 (2009).

29. Stekhoven, D. J. & Bühlmann, P. MissForest—non-parametric missing value imputation for mixed-type data. Bioinformatics 28, 112–118 (2012).

30. Steptoe, A., Deaton, A. & Stone, A. A. Subjective wellbeing, health, and ageing. The Lancet 385, 640–648 (2015).

31. Klein, S. L. & Flanagan, K. L. Sex differences in immune responses. Nat Rev Immunol 16, 626–638 (2016).

32. Dhabhar, F. S. & Mcewen, B. S. Acute Stress Enhances while Chronic Stress Suppresses Cell-Mediated Immunityin Vivo:A Potential Role for Leukocyte Trafficking. *Brain*, Behavior, and Immunity 11, 286–306 (1997).

33. Marshall, G. D. et al. Cytokine Dysregulation Associated with Exam Stress in Healthy Medical Students. *Brain*, Behavior, and Immunity 12, 297–307 (1998).

34. Hamilton, O. S. & Steptoe, A. Socioeconomic determinants of inflammation and neuroendocrine activity: A longitudinal analysis of compositional and contextual effects. Brain, Behavior, and Immunity 107, 276–285 (2022).

35. Garbers, C. et al. Plasticity and cross-talk of Interleukin 6-type cytokines. Cytokine & Growth Factor Reviews 23, 85–97 (2012).

36. Chung, H. Y. et al. Molecular inflammation: Underpinnings of aging and age-related diseases. Ageing Research Reviews 8, 18–30 (2009).

37. Yip, T. et al. Linking discrimination and sleep with biomarker profiles: An investigation in the MIDUS study. Compr Psychoneuroendocrinol 5, 100021 (2020).

38. Rivest, S. Chapter 4 - Interactions Between the Immune and Neuroendocrine Systems. In Progress in Brain Research (ed. Martini, L.) vol. 181 43–53 (Elsevier, 2010).

39. Rajpathak, S. N. et al. Insulin-like growth factor-(IGF)-axis, inflammation, and glucose intolerance among older adults. Growth Horm IGF Res 18, 166–173 (2008).

40. Witkowska-Sędek, E. & Pyrżak, B. Chronic inflammation and the growth hormone/insulin-like growth factor-1 axis. Cent Eur J Immunol 45, 469–475 (2020).

41. Su, S. et al. Genetic and environmental influences on systemic markers of inflammation in middle-aged male twins. Atherosclerosis 200, 213–220 (2008).

42. Sawyers, C. et al. Genetic and environmental influences on cortisol reactivity to a psychosocial stressor in adolescents and young adults. Psychoneuroendocrinology 127, 105195 (2021).

43. Franco, L. et al. Assessment of age-related changes in heritability and IGF-1 gene effect on circulating IGF-1 levels. AGE 36, 9622 (2014).

44. Pankow, J. S. et al. Familial and genetic determinants of systemic markers of inflammation: the NHLBI family heart study. Atherosclerosis 154, 681–689 (2001).

45. Wang, J. & Wang, X. Confirmatory Factor Analysis. in Structural Equation Modeling 29–89 (John Wiley & Sons, Ltd, 2012). doi:10.1002/9781118356258.ch2.

46. Thames, A. D., Irwin, M. R., Breen, E. C. & Cole, S. W. Experienced discrimination and racial differences in leukocyte gene expression. Psychoneuroendocrinology 106, 277–283 (2019).

47. Walker, A. J. et al. Stress, Inflammation, and Cellular Vulnerability during Early Stages of Affective Disorders: Biomarker Strategies and Opportunities for Prevention and Intervention. Frontiers in Psychiatry 5, (2014).

