## Supplemental Materials for "Immune-Neuroendocrine Patterning and Response to Stress. A latent profile analysis in the English Longitudinal Study of Ageing"

**Supplementary Material**

### SM1. Outcomes | Immune and Neuroendocrine Biomarker Assay Procedure for Profiles

**C-reactive Protein.** High-sensitivity plasma CRP (mg/L) was assayed using the N Latex CRP mono Immunoassay on the Behring Nephelometer II analyser (Dade Behring, Milton Keynes, UK). Intra and inter-assay coefficients of variation were <2%. The lower detection limit of the assay was 0.2 mg/L. CRP values >20 mg/L were excluded from analyses (n=116), as these were taken to reflect acute inflammatory processes rather than chronic inflammation.30 CRP was treated as continuous, with higher values indicating greater levels of inflammation.

**Fibrinogen.** Plasma fibrinogen (g/L) was analysed using a modification of the Clauss thrombin clotting method on the Organon Teknika MDA 180 coagulation analyser (Organon Teknika, Durham, USA). Intra and inter-assay coefficients of variation were <7%. The lower detection limit of the assay was 0.5 g/L. Fibrinogen was treated as continuous, with higher values indicating greater levels of inflammation.

**Insulin-like Growth Factor-1.** Serum IGF-1 (nmol/L) was measured using the DPC Immulite 2000 method, by an electrochemiluminescent immunoassay on IDS ISYS Analyser. Inter and intra-assay coefficients of variation were <14%. IGF-1 was treated as continuous, with lower values indicating greater neuroendocrine activity.

**Hair Cortisol.** Hair strands ~3cm, weighing ~10mg were collected from the posterior vertex, as close to the scalp as possible. Assuming an average hair growth of ∼1cm per month,[16] the hair segment closest to the scalp is thought to provide a measure of the average cortisol output over the preceding three months prior to sampling. Exclusion criteria for hair sampling included pregnancy, breastfeeding, select scalp conditions, having <2cm of hair length, and an inability keep head still. Hair cortisol concentrations were analysed at the Technische Universität Dresden (Germany) in two separate phases (2015 and 2018) owing to financial constraints. Cortisol levels were assayed using high performance liquid chromatography-mass spectrometry (LC/MS) following a standard wash and steroid extraction procedure,[17] and were expressed in pg/mg. Data was log-transformed, as the distribution was positively skewed.

**SM2. Covariates | PGS for Immune and Neuroendocrine Biomarkers Construction**

**Genetic Data.** The genome-wide genotyping was performed at University College London (UCL) Genomics in 2013-2014 with the funding the Economic and Social Research Council (ESRC) using the llumina HumanOmni2.5 BeadChips (HumanOmni2.5-4v1, HumanOmni2.5-8v1.3), which measures ~2.5 million markers that capture the genomic variation down to 2.5% minor allele frequency.

**Quality Control.** The methods employed for quality control of genomic data in the ELSA study are those outlined by the Health and Retirement Study (HRS).[1] This was done to harmonise the research across the age-related longitudinal studies by adopting a consistent methodology. Single-nucleotide polymorphism (SNPs) were excluded if they were non-autosomal, Minor allele frequency (MAF) was <1%, if more than 2% of genotype data were missing and if the Hardy-Weinberg Equilibrium $p<10^{-4}$. Samples were removed based on call rate (<0.99), heterozygosity, relatedness and if the recorded sex phenotype was inconsistent with genetic sex. To identify ancestrally homogenous analytic samples the ELSA genomic samples use a combination of both self-reported ethnicity and analyses of genetic ancestry. To improve genome coverage, we imputed untyped quality-controlled genotypes to the Haplotype Reference Consortium[2] using the University of Michigan Imputation Server.[3] Post-imputation, we kept variants that were genotyped or imputed at INFO>0.80, in low linkage disequilibrium ($R^2<0.1$) and with Hardy-Weinberg Equilibrium $p$-value$>10^{-5}$. After the sample quality control 7179780 variants were retained for further analyses. To account for potentially biasing ancestry differences in genetic structures, a principal components (PCs) analysis was conducted, retaining the top 10 PCs,[4] which were subsequently used to adjust for possible population stratification in the association analyses.[4,5] Genetic ancestry was estimated via comparison of participants' genotypes to global reference populations using principal component analyses (PCA). Because PCA allows examining population structure in a cohort by determining the average genome-wide genetic similarities of individual samples, derived principal components (PCs) can be used to group individuals with shared genetic ancestry, to identify outliers, and as covariates, to reduce false positives due to population stratification. Although up to 98% of the ELSA participants self-described to be of European cultural background, PC highlighted the present of ancestral admixture in n=65 (0.9%) individuals (implying these individuals had ancestors from two or more populations. Even though this type of labelling of ancestral populations oversimplifies the complexity of human genetic variation, accounting for systematic differences in allele frequencies is necessary for genetic analyses. Therefore, these participants with ancestral admixture were removed from the analyses. The final sample includes all self-reported European participants that had PC loadings within ± one standard deviations of the mean for eigenvectors one. PCs were then re-calculated to further account for population stratification. Therefore, our analytic sample included the full ELSA sample that provided genetic samples and passed quality control. We further utilized the PCs for adjusting for possible population stratification in the association analyses.

**Polygenic risk scores (PGS).** PGS for C-reactive protein in ELSA was contracted using the results from two genome-wide association studies (GWAS), based on both HapMap and 1000 Genomes imputed data and encompassing data from 88 studies comprising 204,402 European individuals.[6] The GWAS meta-analyses of C-reactive protein revealed 58 distinct genetic loci ($p < 5 \times 10^{-8}$). After adjustment for body mass index in the regression analysis, the associations at all except three loci remained. The lead variants at the distinct loci explained up to 7.0% of the variance in circulating amounts of C-reactive protein. Further 66 gene sets that were organized in two substantially correlated clusters were identified, one mainly composed of immune pathways and the other characterized by metabolic pathways in the liver. The GWAS summary statistics for this phenotype contained 10,019,203 SNPs; of these, 1,301,076 SNPs overlapped with the ELSA genetic database and were included in the PGS for C-reactive protein phenotype. PGS for IGF-1 was calculated using summary statistics from GWAS that included 10280 men and women in the analyses, comprising 1712 participants in the Cardiovascular Health Study (CHS), 3507 in the Framingham Heart Study (FHS), 1607 participants in the Cooperative Research in the Region of Augsburg (KORA) study and 3454 in the Study of Health in Pomerania (SHIP). Analyses of SNP associated with IGF-1 concentrations revealed that rs700752 was associated with IGF-I concentrations ($p = 4.9 \times 10^{-9}$), but this was attenuated (meta-analysis $p = 0.038$) after adjustment for IGFBP-3 concentrations. Three additional SNPs achieved $p < 10^{-6}$ in relation to IGF-I concentrations: rs2153960 on chromosome 6q21, MAF = 0.31, $p = 5.1 \times 10^{-7}$; rs1245541 on chromosome 10q22.1, MAF = 0.39, $p = 5.0 \times 10^{-7}$; rs7780564 on chromosome 7p21.3, MAF = 0.45, $P = 3.9 \times 10^{-7}$. PGS for Morning Plasma Cortisol in ELSA was contracted using the results from the CORtisol NETwork (CORNET) consortium, which undertook the GWAS meta-analysis for plasma cortisol in 12,597 White participants from 11 western European population-based cohorts and replicated their results in 2,795 participants from three independent cohorts.[7] Cortisol was measured by immunoassay in blood samples collected from study participants between 07:00h and 11:00h. Each study performed single marker association tests, and study-specific linear regression models which used z-scores of log-transformed cortisol, additive SNP effects, and were adjusted for age and sex (model 1); age, sex, and smoking (model 2); or age, sex, smoking and body mass index (model 3). Imputation of the gene-chip results used the HapMap CEU population, build 36. The results indicate that <1% of variance in plasma cortisol is accounted for by genetic variation in a single region of chromosome 14. The CORNET GWAS summary statistics for this phenotype contained 2,660,191 SNPs; of these, 837,709 SNPs overlapped with the ELSA genetic database and were included in the PGS for Morning Plasma Cortisol phenotype.

**SM3. Statistical Procedure and Parameters for Latent Profile Analysis**

An LPA model for observed variable *A* can be expressed as:

$$\sigma\frac{2}{A} = \sum_{t=1}^{T} \pi_t(\mu_{At} - \mu_A)^{\ 2} + \sum_{t=1}^{T} \pi_t \sigma_{At}^2$$

where $\mu_{At}$ and $\sigma_{At}^2$ denote (*t*) class-specific means and variances for variable *A*, and $\pi_t$ show the proportion of *N* participants that belong to class *t*. The number of latent profiles was determined on the basis of the Akaike information criterion (AIC),[11] Bayesian information criterion (BIC),[12] and adjusted Bayesian information criterion (aBIC).[13] The information criteria and the likelihood ratio tests indicated the goodness of fit of different latent profile models, with the best model being the one with the lowest AIC, BIC, and aBIC values. The entropy statistic that provides the quality of the classification model, and the average posterior probabilities for each latent profile, indicating profile membership classification errors, were also taken into account.[14] The closer to 1 these indicators were, the better the classification quality.[15] A common cut-off point for posterior probabilities is 0.70 or above.[16] An entropy of 0.80 or greater indicates clear profile separation.[17] Every profile must contain more than 5% of participants and the profiles must be of good theoretical interpretability.[18]

*Figure S1.* **Directed acyclic graph (DAG) conceptually representing associations between study variables**

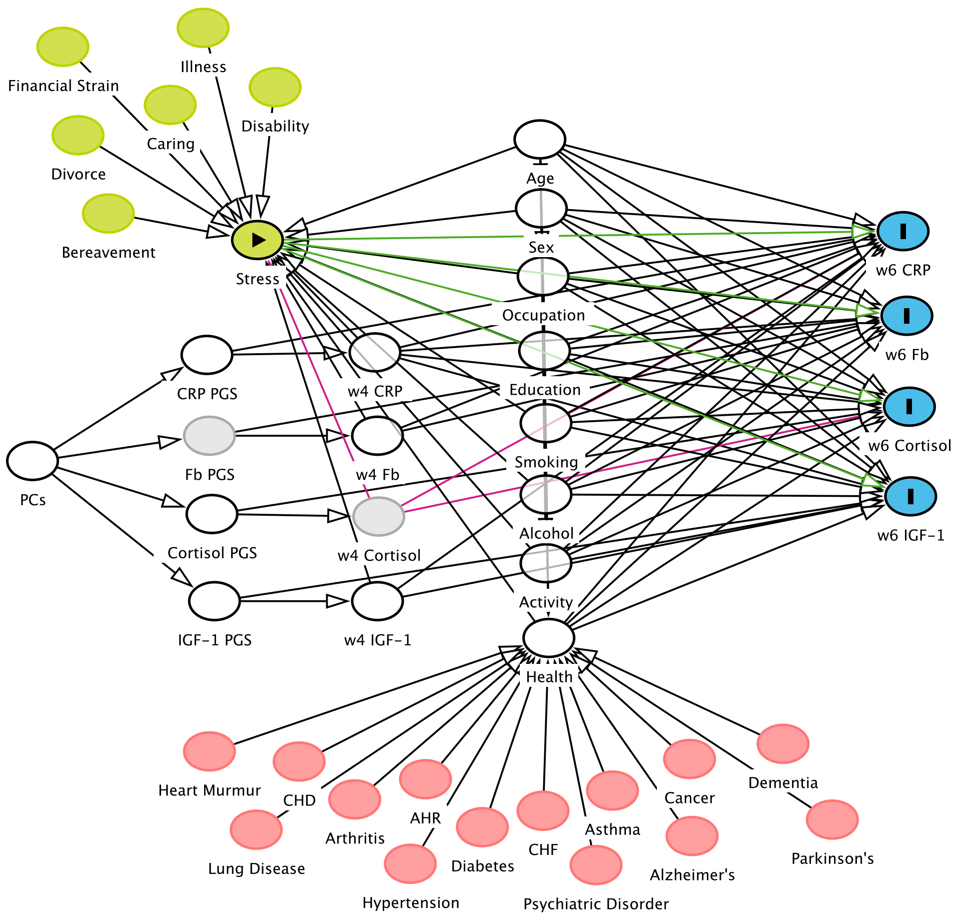


**KEY**

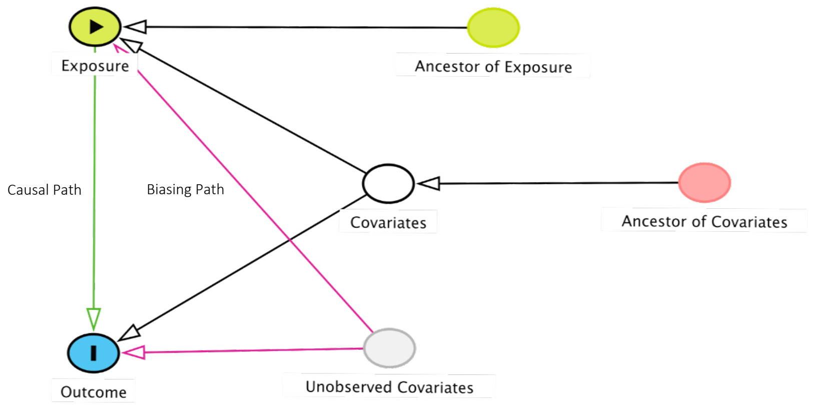

*Figure S2.* **The percentage of participants belonging to each immune and neuroendocrine biomarker profile with 95% confidence intervals for the wave 4 three-profile solution (N = 4,934)**

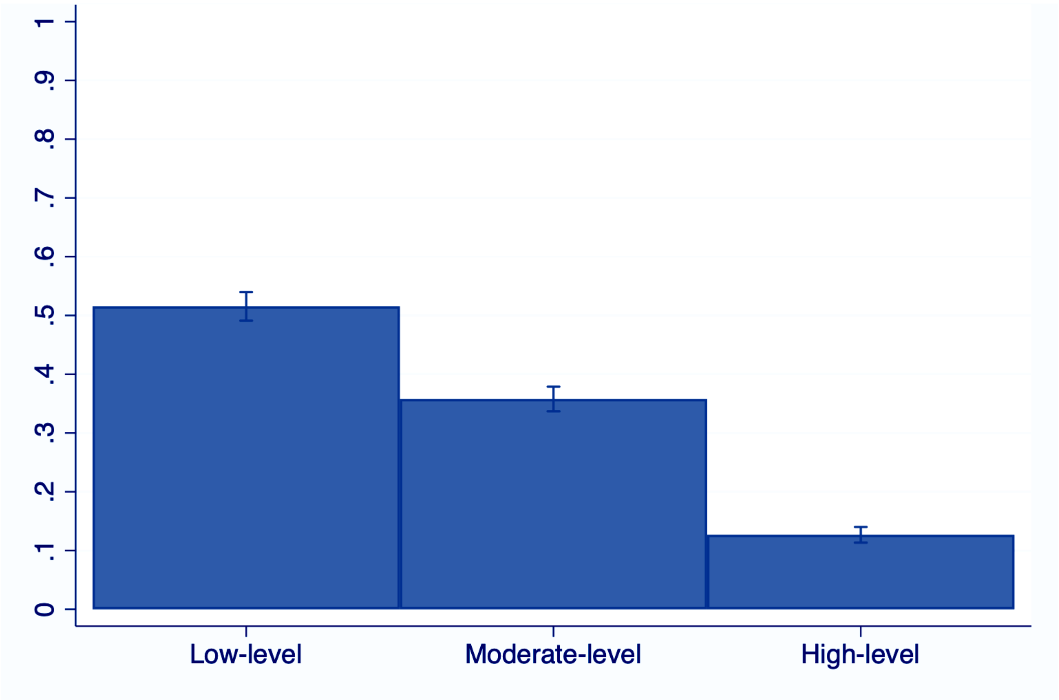


| **Profile** | **N** | **%** |
|---|---|---|
| **1** | 2,560 | 51.88 |
| **2** | 1776 | 36.00 |
| **3** | 598 | 12.12 |

*Figure S3.* **The predicted mean values of immune and neuroendocrine biomarker profiles for the wave 4 three-profile solution (N = 4,934)**

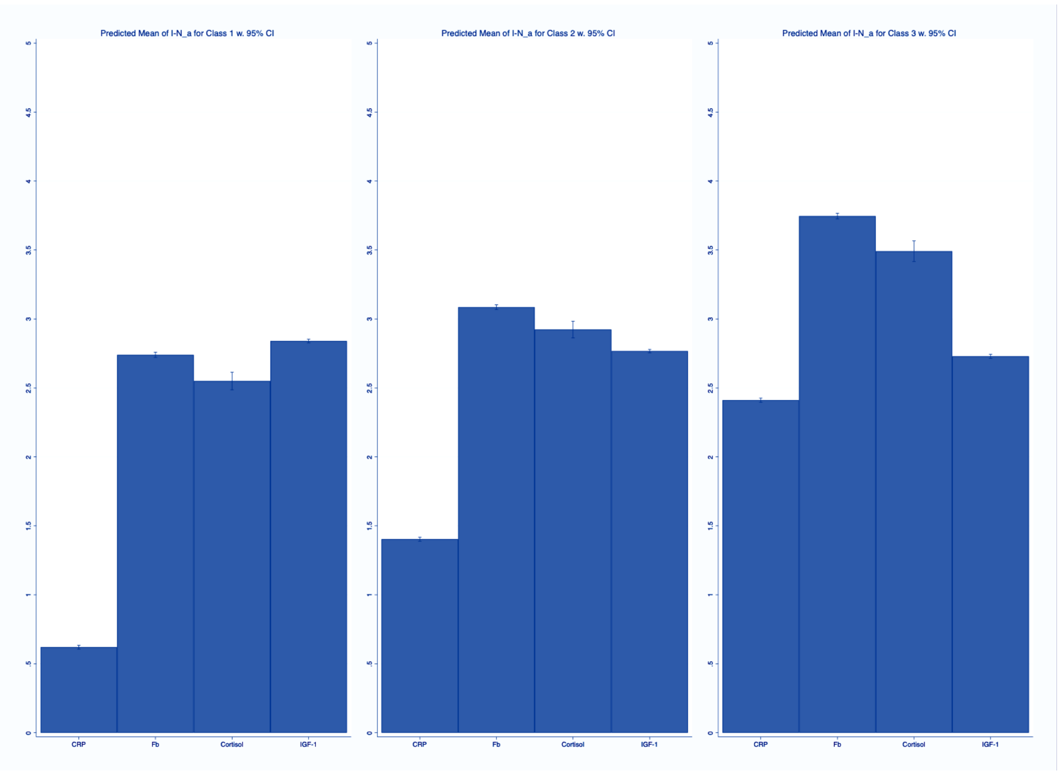

*Figure S4.* **Flow chart of missingness and the analytic sample for imputed data**

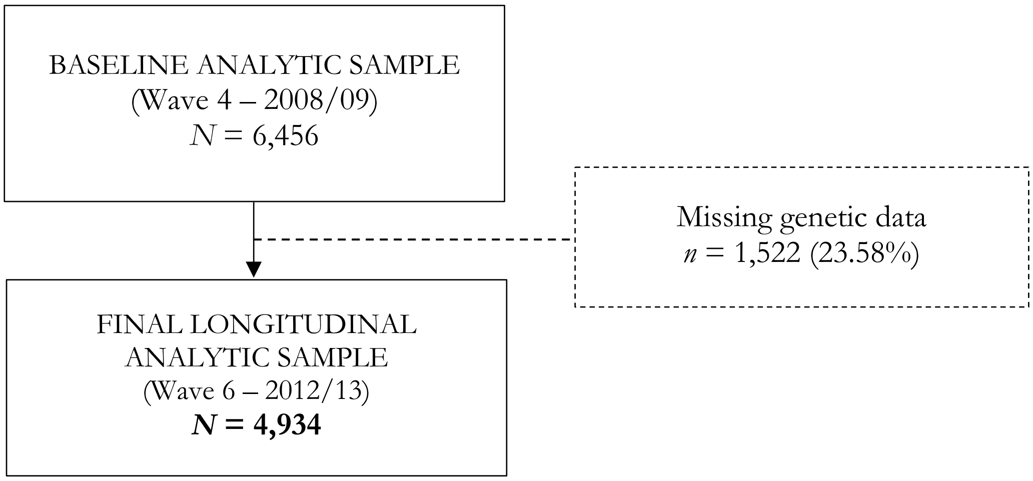

***Figure S5.*** **Flow chart of missingness and the analytic sample for complete cases**

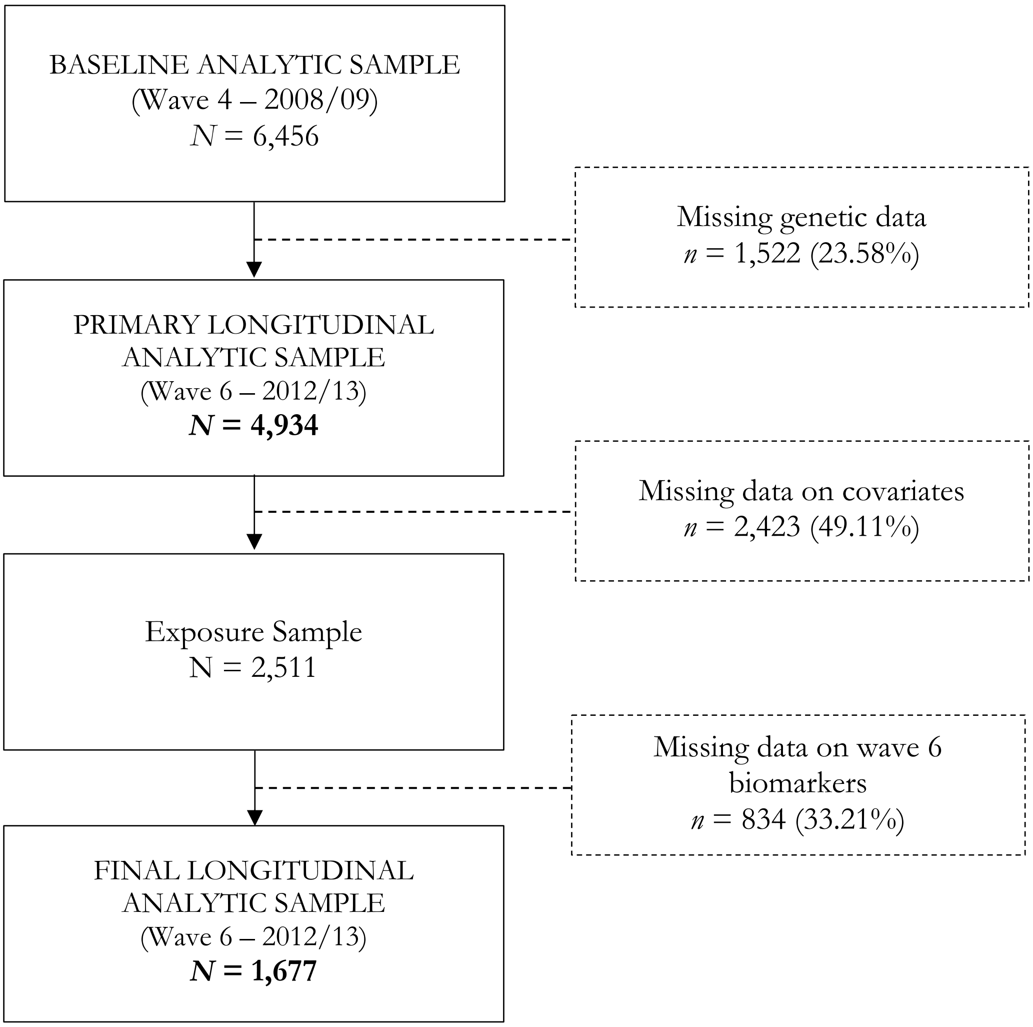

*Figure S6.* **Percentage of total stress experienced (N=8,083)**

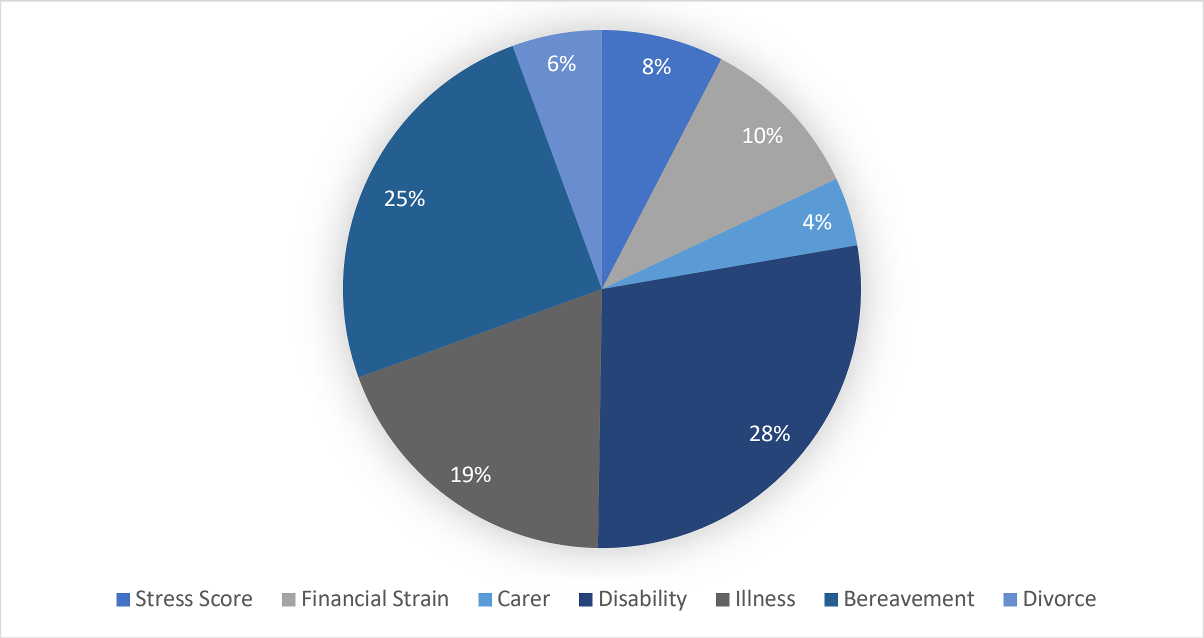

*Figure S7.* **Independent stress experiences of the sample**

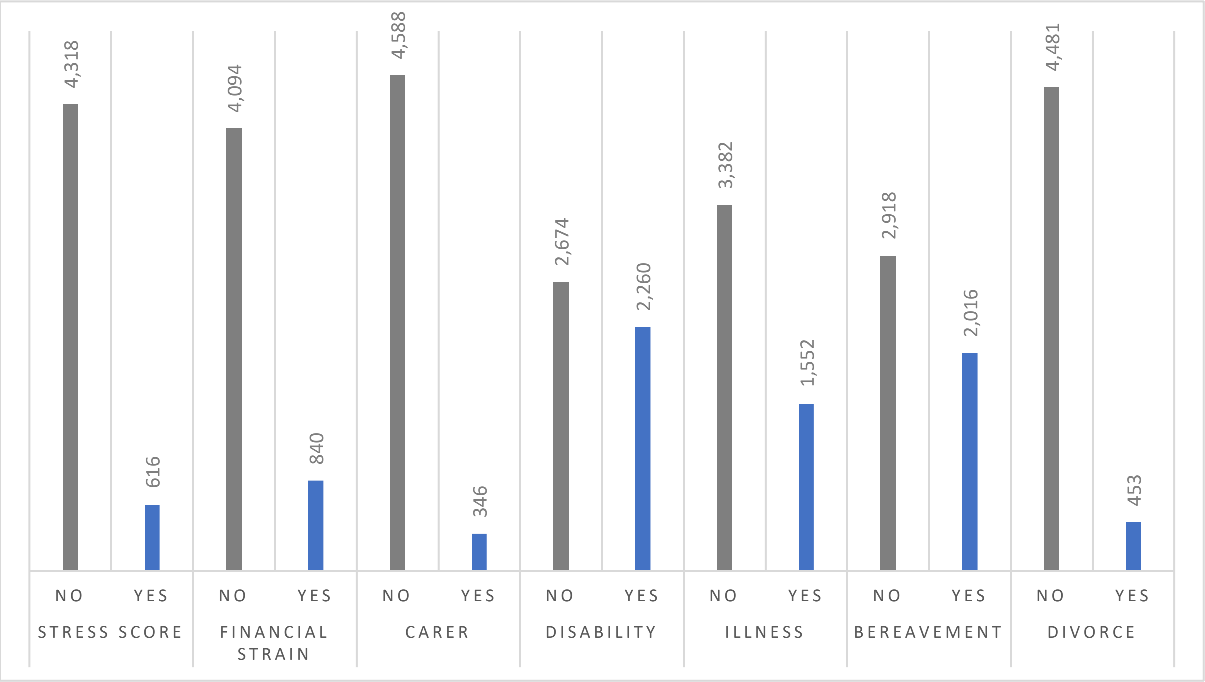

*Figure S8* [a-g]. Mean immune and neuroendocrine biomarker levels for a one to seven profile solution (N = 4,934)

a

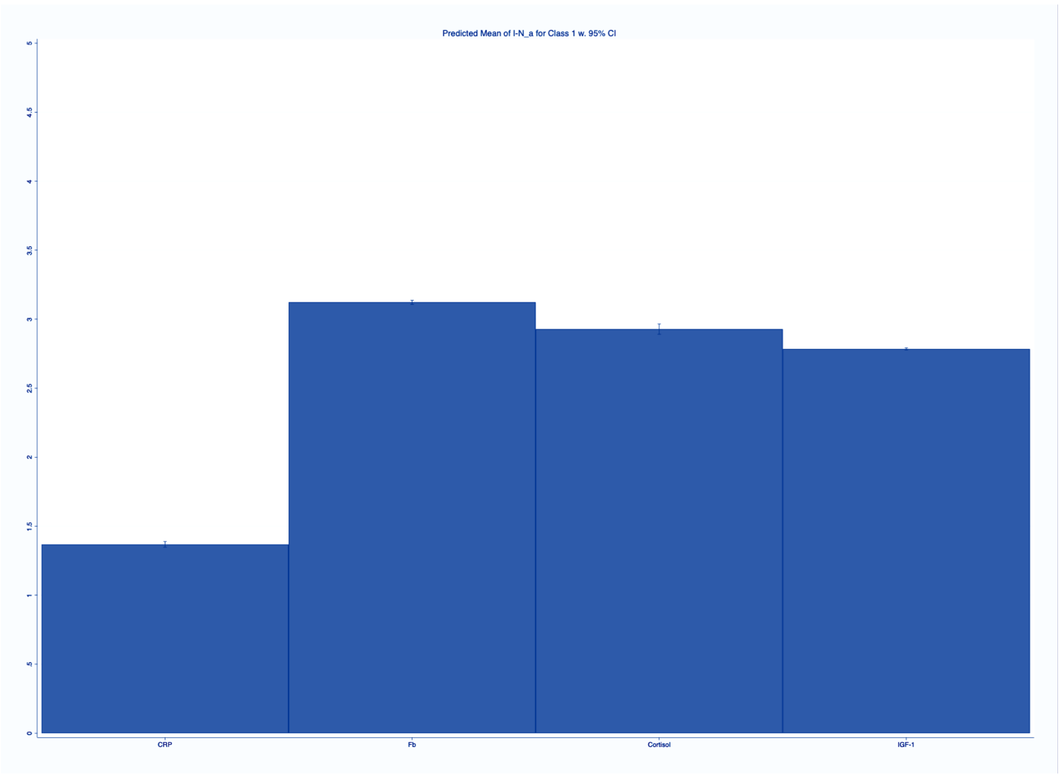


b

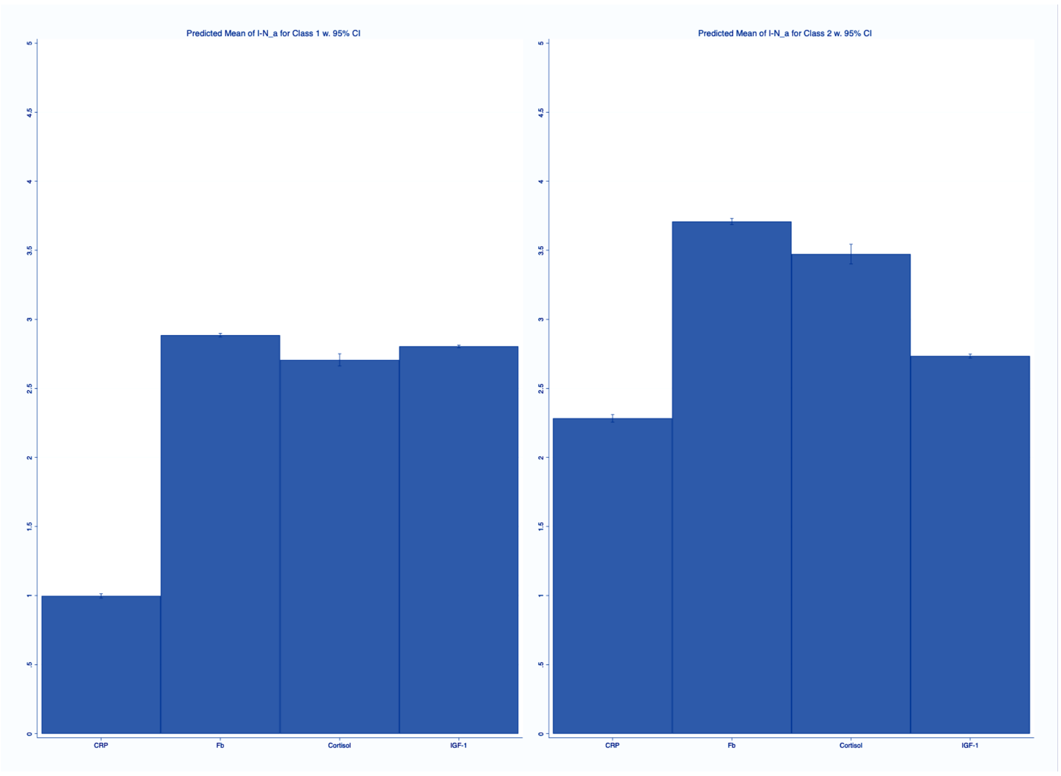

c

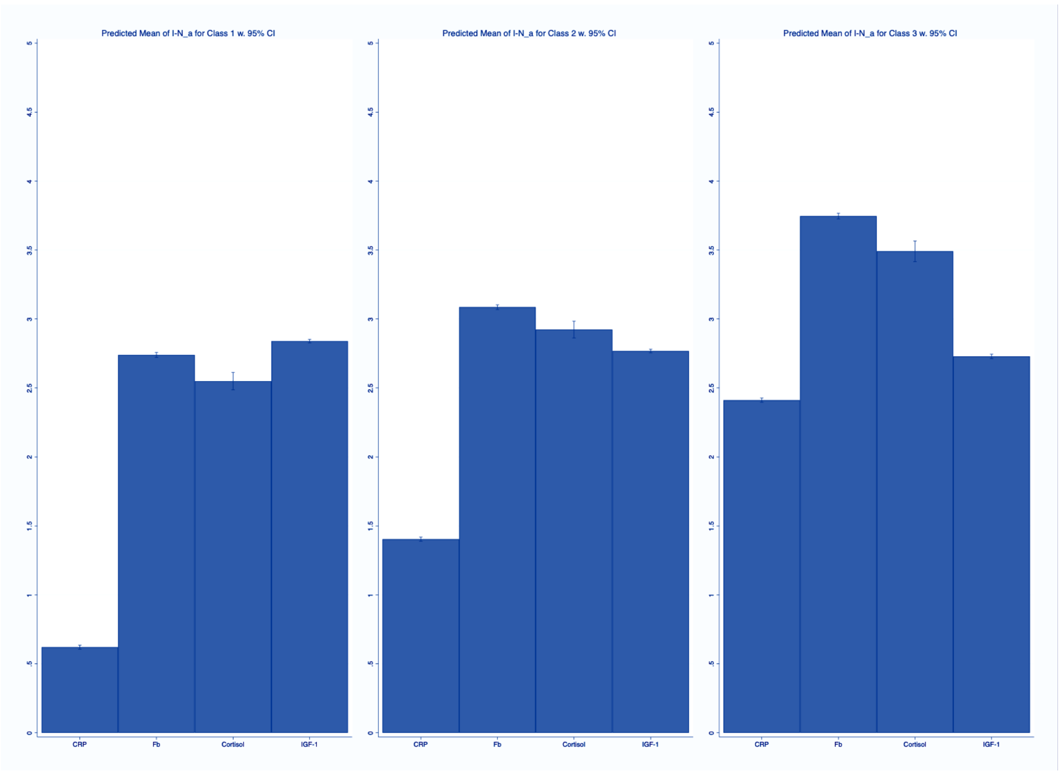
Predicted Mean of I-N_a for Class 1 w. 95% CI
Predicted Mean of I-N_a for Class 2 w. 95% CI
Predicted Mean of I-N_a for Class 3 w. 95% CI
0
.5
1
1.5
2
2.5
3
3.5
4
4.5
5
CRP
Fb
Cortisol
IGF-1

d

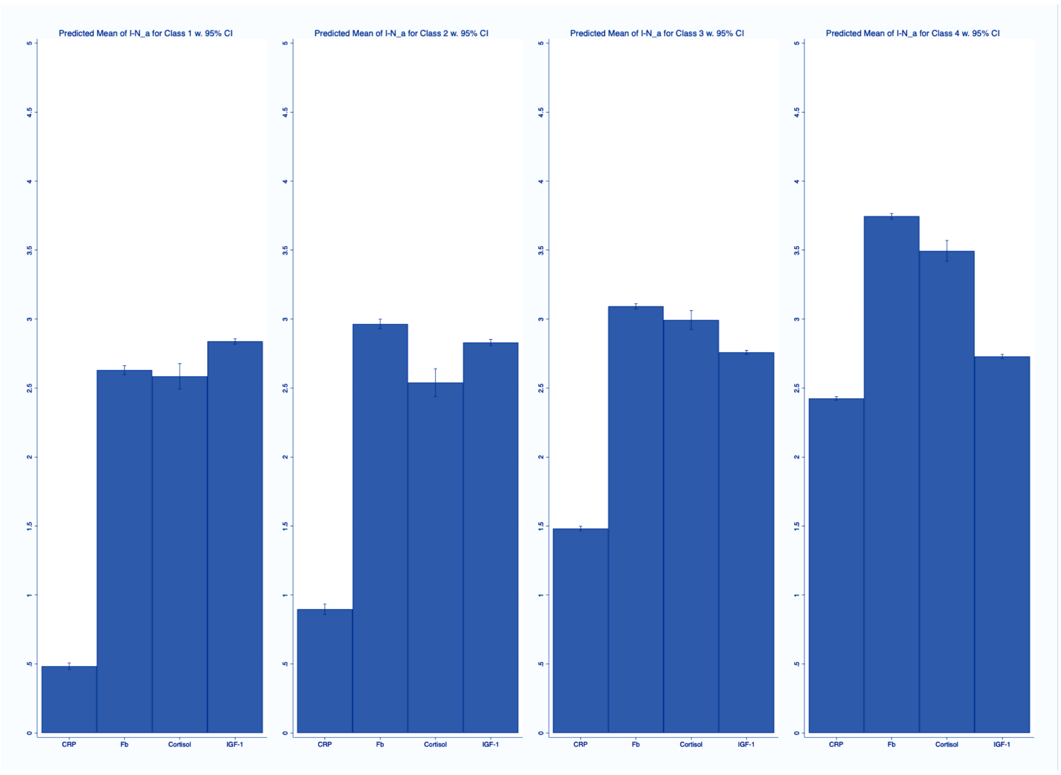
Predicted Mean of I-N_a for Class 1 w. 95% CI
Predicted Mean of I-N_a for Class 2 w. 95% CI
Predicted Mean of I-N_a for Class 3 w. 95% CI
Predicted Mean of I-N_a for Class 4 w. 95% CI
0
.5
1
1.5
2
2.5
3
3.5
4
4.5
5
CRP
Fb
Cortisol
IGF-1

e

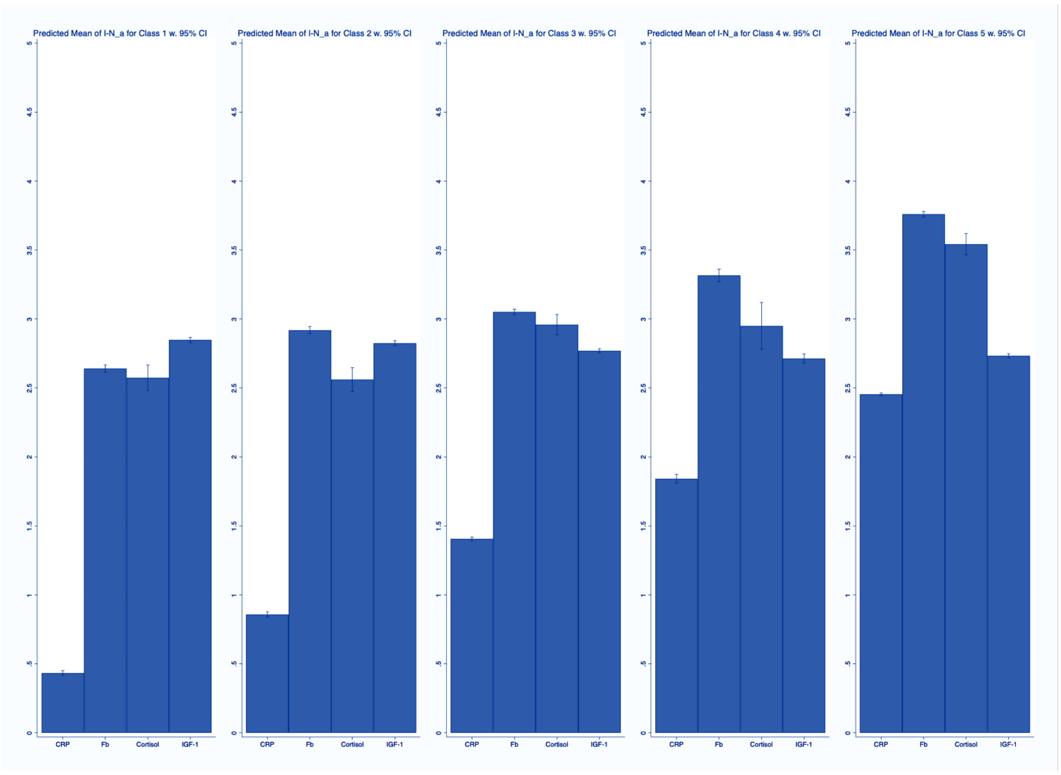
Predicted Mean of I-N_a for Class 1 w. 95% CI
Predicted Mean of I-N_a for Class 2 w. 95% CI
Predicted Mean of I-N_a for Class 3 w. 95% CI
Predicted Mean of I-N_a for Class 4 w. 95% CI
Predicted Mean of I-N_a for Class 5 w. 95% CI
0
.5
1
1.5
2
2.5
3
3.5
4
4.5
5
CRP
Fb
Cortisol
IGF-1

f

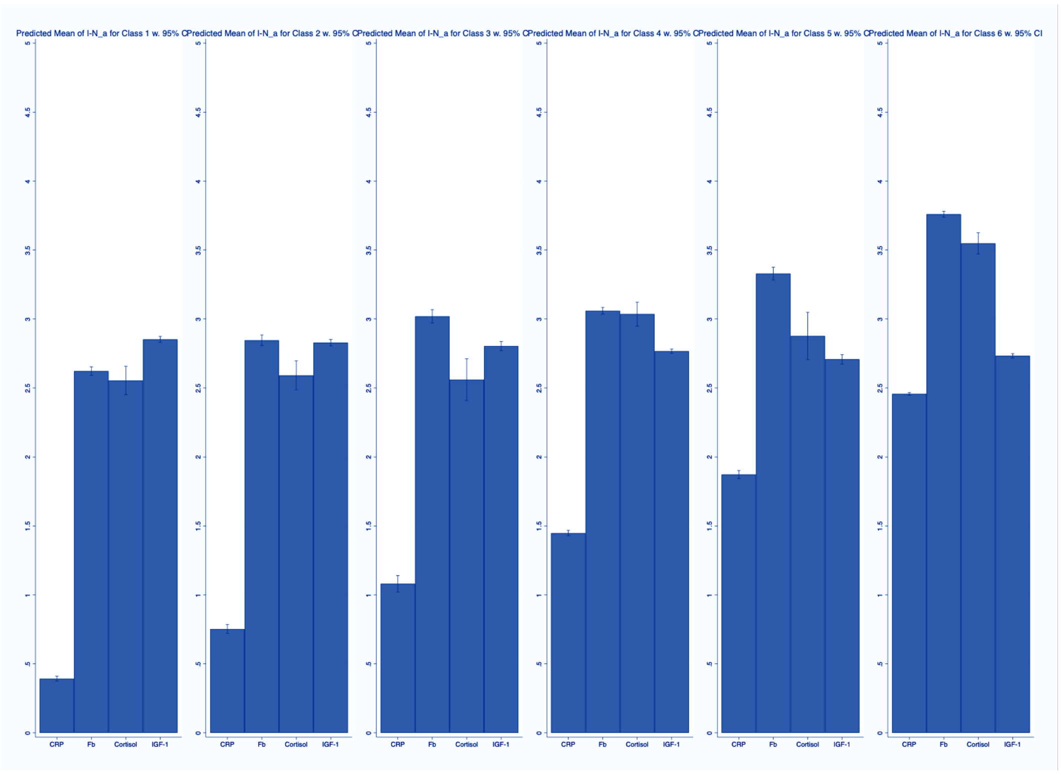
Predicted Mean of I-N_a for Class 1 w. 95% C
Predicted Mean of I-N_a for Class 2 w. 95% C
Predicted Mean of I-N_a for Class 3 w. 95% C
Predicted Mean of I-N_a for Class 4 w. 95% C
Predicted Mean of I-N_a for Class 5 w. 95% C
Predicted Mean of I-N_a for Class 6 w. 95% CI
0
.5
1
1.5
2
2.5
3
3.5
4
4.5
5
CRP
Fb
Cortisol
IGF-1

g

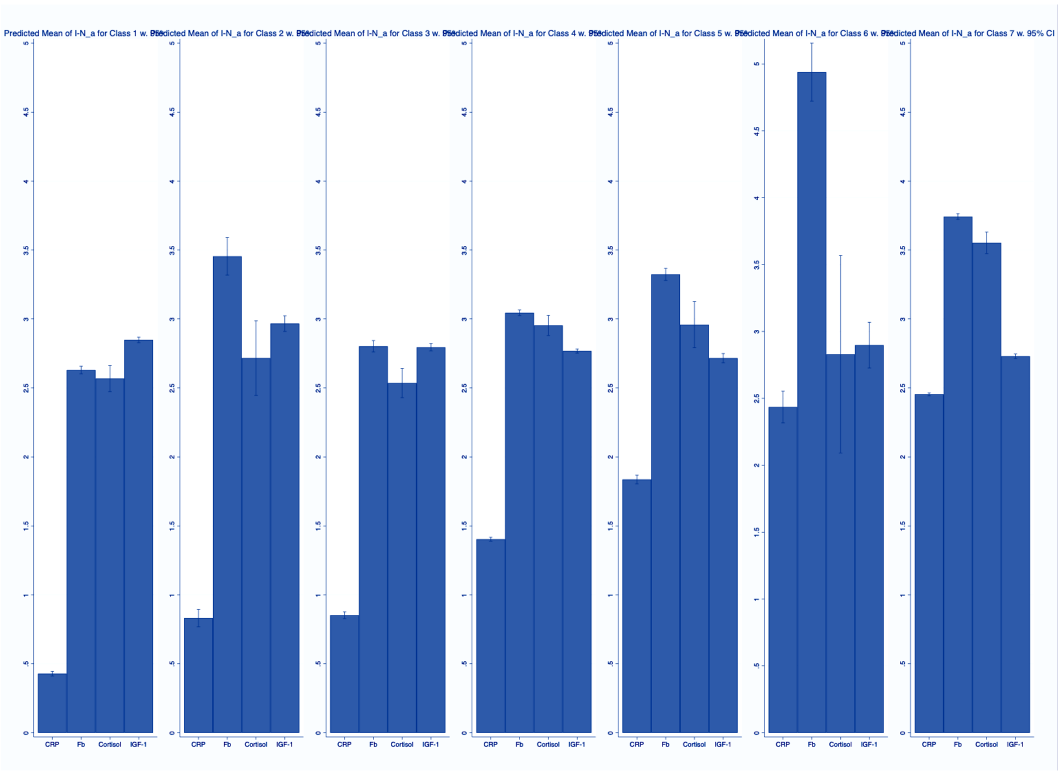
Predicted Mean of I-N_a for Class 1 w. 95% CI
Predicted Mean of I-N_a for Class 2 w. 95% CI
Predicted Mean of I-N_a for Class 3 w. 95% CI
Predicted Mean of I-N_a for Class 4 w. 95% CI
Predicted Mean of I-N_a for Class 5 w. 95% CI
Predicted Mean of I-N_a for Class 6 w. 95% CI
Predicted Mean of I-N_a for Class 7 w. 95% CI
CRP
Fb
Cortisol
IGF-1

***Figure S9.*** **The Akaike information criterion (AIC), Bayesian information criterion (BIC) for the seven profile LPA model fit**

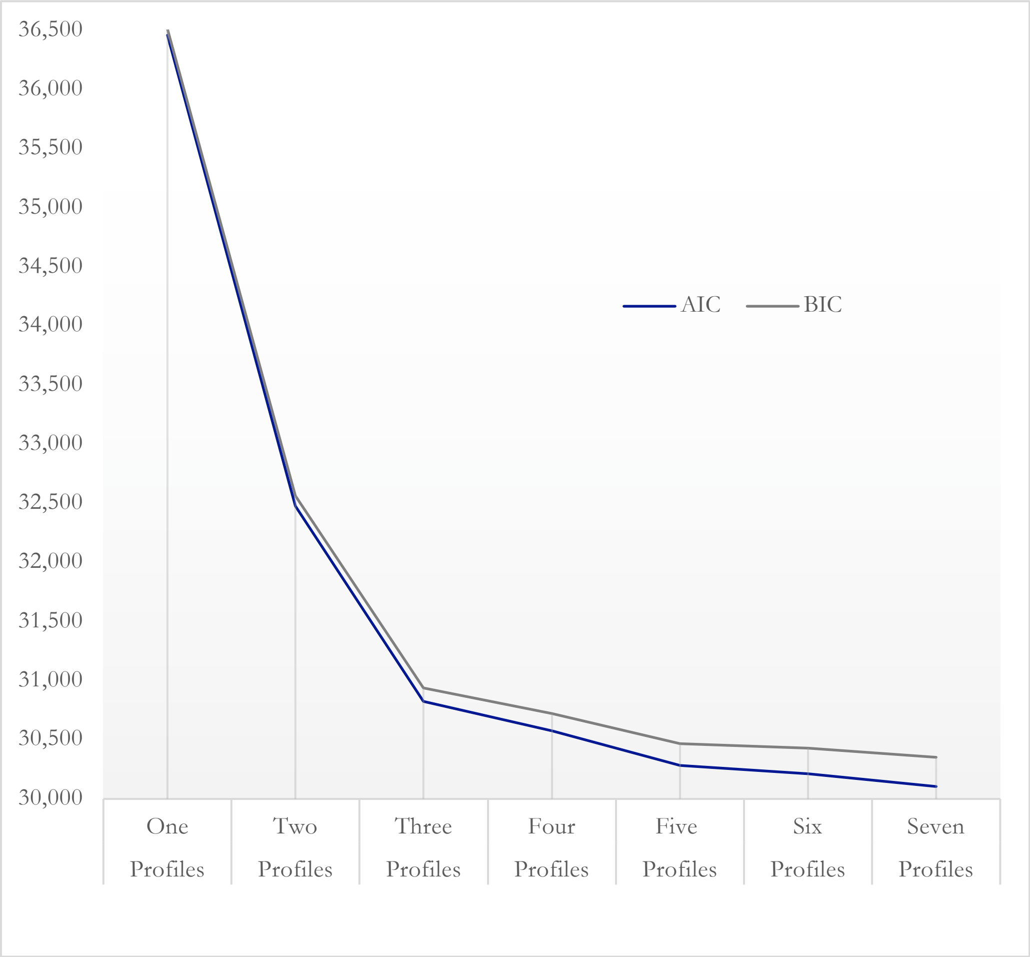

*Figure S10.* **The entropy and normalised entropy statistic for the seven profile LPA model fit**

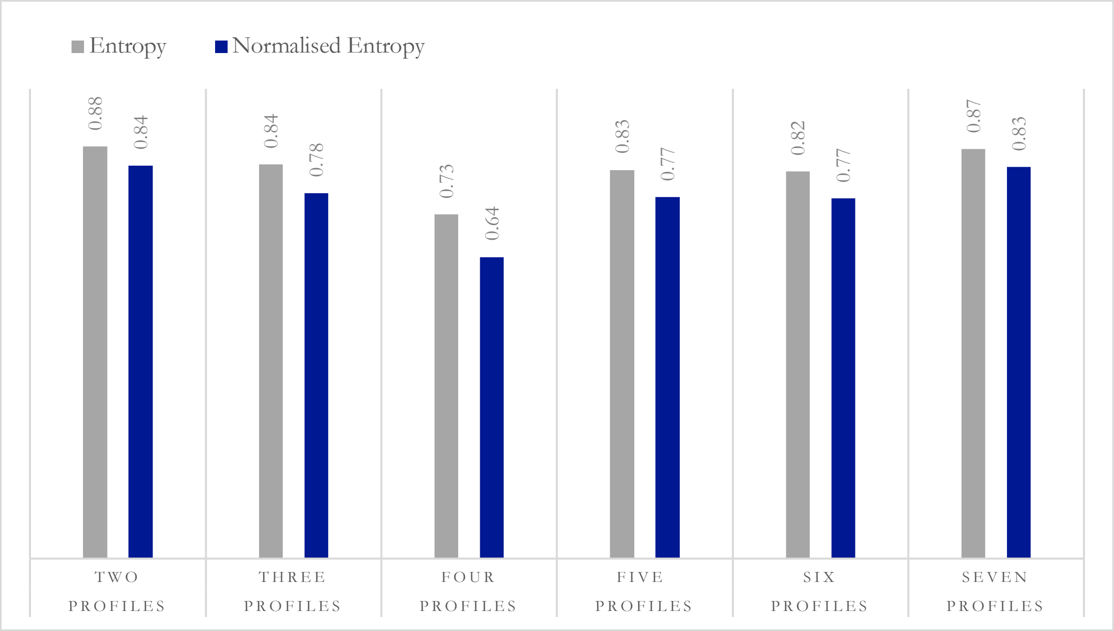

*Figure S11.* **The mean posterior probabilities for the seven profile LPA model fit**

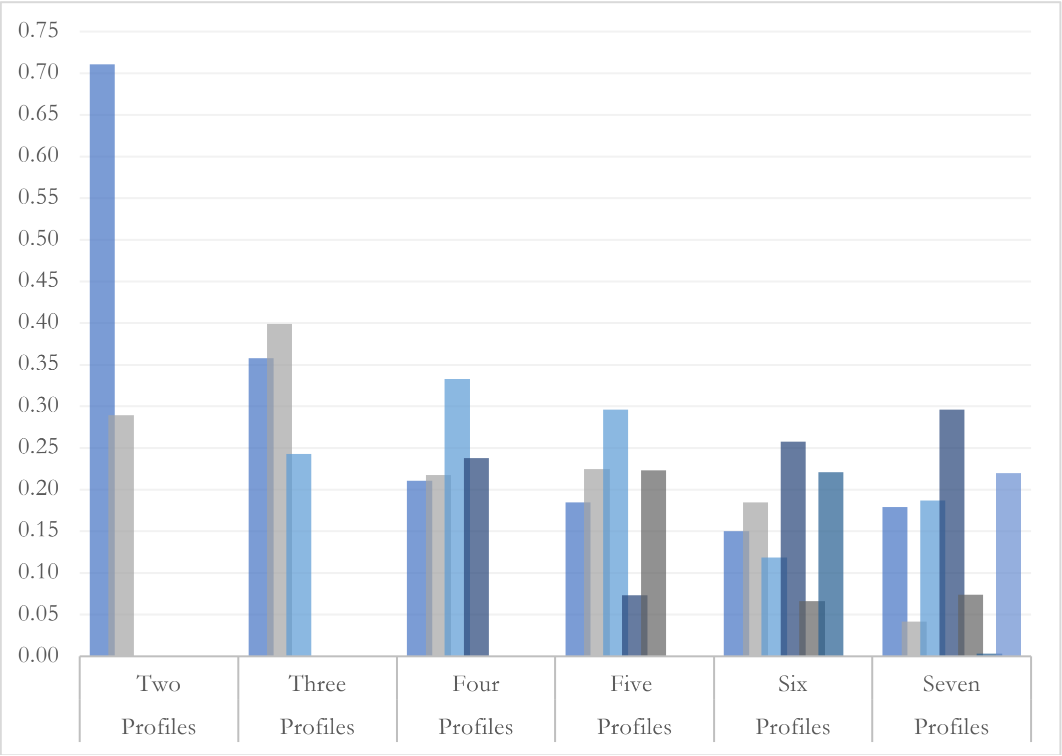

*Figure S12.* **The percentage of participants belonging to each immune and neuroendocrine profile**

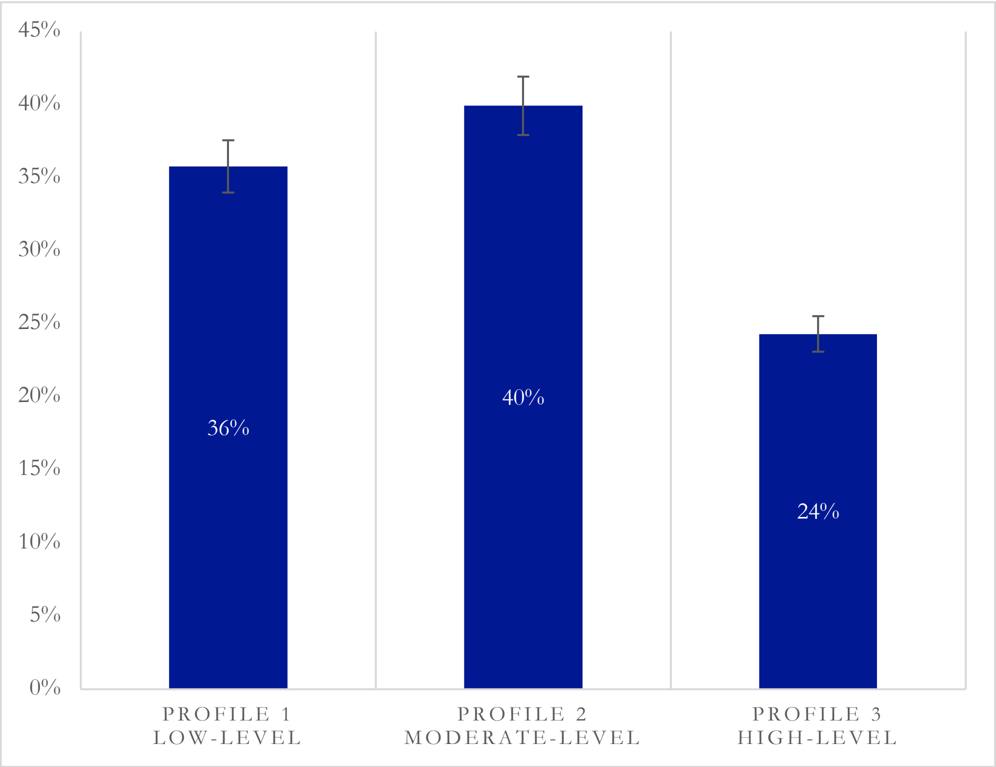

*Figure S13.* **The predicted mean values of immune and neuroendocrine biomarker profiles for the three-profile solution using complete case data (N = 1,677)**

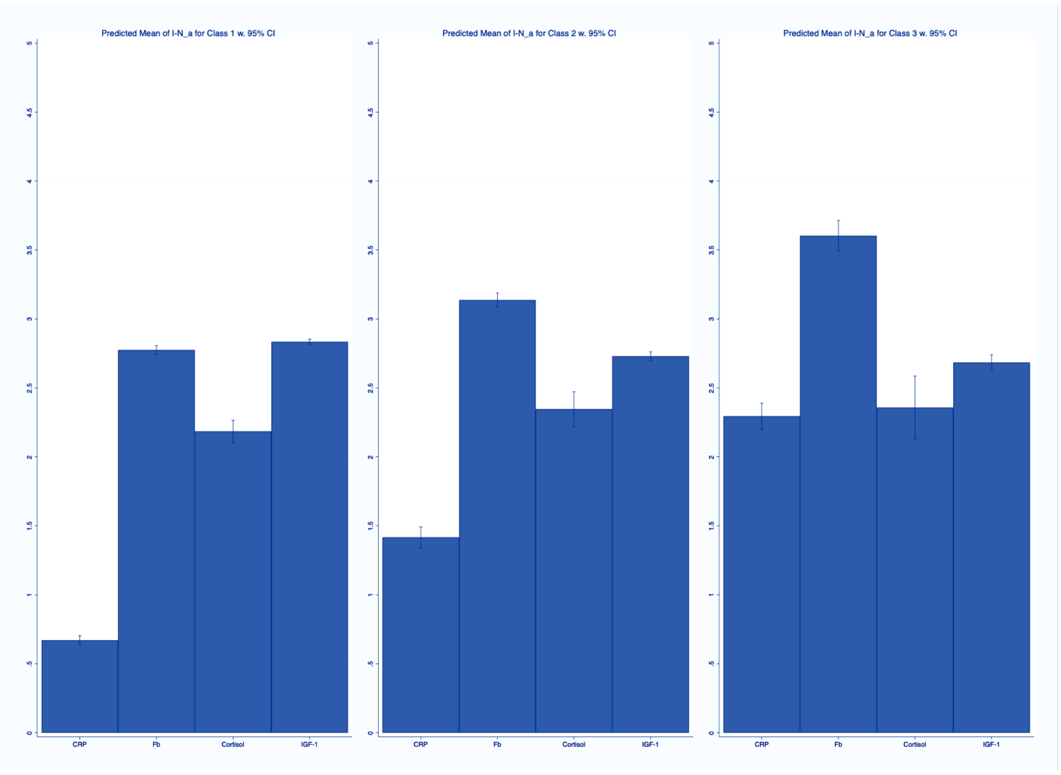

*Figure S14.* **The percentage of participants belonging to each immune and neuroendocrine biomarker profile with 95% confidence intervals for the three-profile solution using complete case data (N = 1,677)**

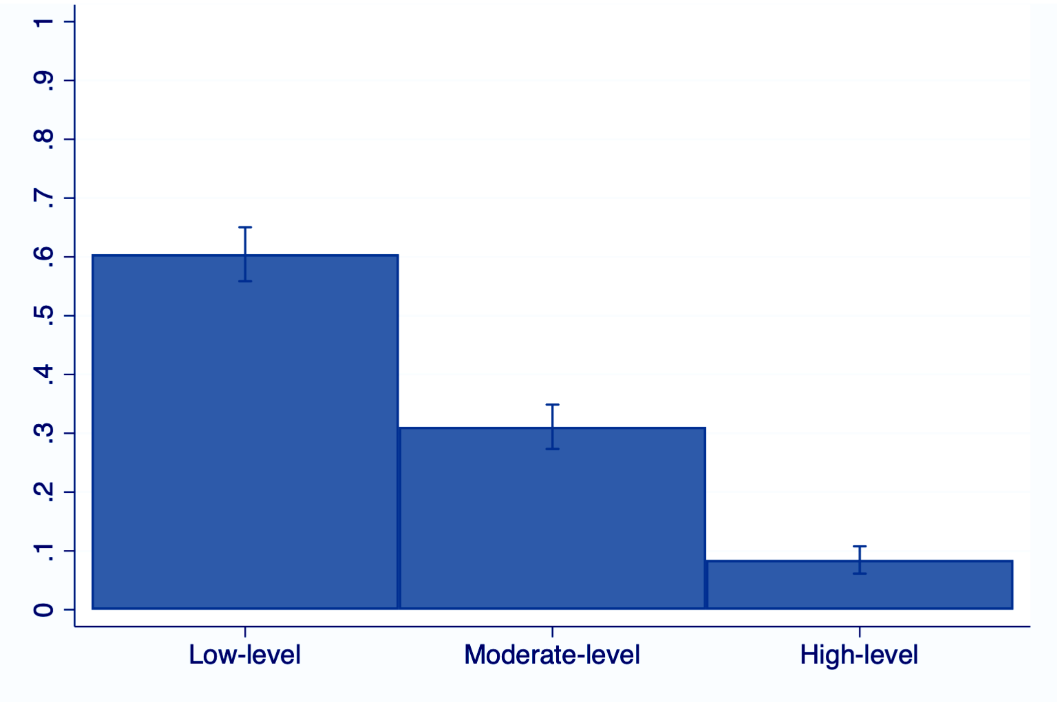


| Profile | N | % |
|---|---|---|
| **1** | 1,028 | 61.30 |
| **2** | 517 | 30.83 |
| **3** | 132 | 7.87 |

*Table S1.* **A comparison of core, imputed, and observed sample characteristics, with missing data**

| Variable | | Missing Data | Core Sample (N=6,512) | | Imputed (N=4,934) | | Complete Cases (N=1,677) | |
|---|---|---|---|---|---|---|---|---|
| | | N \| % | N / Mean (SD) | % / Range | N / Mean (SD) | % / Range | N / Mean (SD) | % / Range |
| Age | | 0 \| 0 | 65.40 (9.42) | 50-99 | 66.31 (9.35) | 50-99 | 65.04 (8.01) | 50-99 |
| Age (Binary) | < Median | 0 \| 0 | 3,684 | 56.57 | 2,437 | 49.39 | 826 | 49.25 |
| | ≥ Median | | 2,828 | 43.43 | 2,497 | 50.61 | 851 | 50.75 |
| Sex | Male | 0 \| 0 | 2,941 | 45.16 | 2,235 | 45.30 | 540 | 32.20 |
| | Female | | 3,571 | 54.84 | 2,699 | 54.70 | 1,137 | 67.80 |
| Education | Higher | 15 \| 0.23 | 2,111 | 32.42 | 1,585 | 32.12 | 567 | 33.81 |
| | Primary/Secondary/Tertiary | | 2,075 | 31.86 | 1,544 | 31.29 | 562 | 33.51 |
| | Alternative or None | | 2,326 | 35.72 | 1,805 | 36.58 | 548 | 32.68 |
| Occupational Social Class | Managerial/Professional | 149 \| 2.31 | 2,430 | 37.32 | 1,790 | 36.28 | 598 | 35.66 |
| | Intermediate Occupations | | 1,639 | 25.17 | 1,264 | 25.62 | 455 | 27.13 |
| | Routine/Manual | | 2,443 | 37.52 | 1,880 | 38.10 | 624 | 37.21 |
| Smoking Status | Non-smokers/Ex-smokers | 44 \| 1.68 | 5,663 | 86.96 | 4,306 | 87.27 | 1,485 | 88.55 |
| | Smokers | | 849 | 13.04 | 628 | 12.73 | 192 | 11.45 |
| Alcohol Consumption | <3 days a week | 548 \| 8.49 | 4,253 | 65.31 | 3,171 | 64.27 | 1,029 | 61.36 |
| | ≥3 days a week | | 2,259 | 34.69 | 1,763 | 35.73 | 648 | 38.64 |
| Physical Activity | Moderately/Vigorously Active | 34 \| 0.53 | 1,781 | 27.35 | 1,338 | 27.12 | 383 | 22.84 |
| | Sedentary | | 4,731 | 72.65 | 3,596 | 72.88 | 1,294 | 77.16 |
| PGS for CRP | Low | 1,522 \| 23.57 | 3,945 | 79.96 | 3,945 | 79.96 | 1,353 | 80.68 |
| | High | | 989 | 20.04 | 989 | 20.04 | 324 | 19.32 |
| PGS for Cortisol | Low | 1,522 \| 23.57 | 3,969 | 80.44 | 3,969 | 80.44 | 1,377 | 82.11 |
| | High | | 965 | 19.56 | 965 | 19.56 | 300 | 17.89 |
| PGS for IGF-1 | Low | 1,522 \| 23.57 | 3,929 | 79.63 | 3,929 | 79.63 | 1,345 | 80.20 |
| | High | | 1,005 | 20.37 | 1,005 | 20.37 | 332 | 19.80 |
| Stress Score | | 0 \| 0 | 1.56 (.90) | 0-6 | 1.51(.90) | 0-6 | 1.50 (.89) | 0-6 |
| Binary Stress Score | No | 0 \| 0 | 5,655 | 86.84 | 4,318 | 87.52 | 1,487 | 88.67 |
| | Yes | | 857 | 13.16 | 616 | 12.48 | 190 | 11.33 |
| CRP* (mg/L; Baseline) | | 159 \| 2.46 | 1.18 (.68) | .18-3.05 | 1.19 (.68) | .18-3.04 | 1.13 (.65) | .18–3.03 |
| CRP* (mg/L; Follow-up) | | 2,374 \| 36.77 | 1.35 (.71) | .10-3.05 | 1.37 (.73) | .10-3.05 | 1.04 (.60) | .10-3.03 |
| Fibrinogen (g/L; Baseline) | | 188 \| 2.91 | 3.37 (.56) | 1.30-5.90 | 3.38 (.56) | 1.30–5.90 | 3.32 (.52) | 1.70–5.30 |
| Fibrinogen (g/L; Follow-up) | | 2,373 \| 36.76 | 3.11 (.51) | 1.30-5.80 | 3.12 (.54) | 1.50–5.80 | 2.96 (.50) | 1.60–5.20 |
| Cortisol* (pg/mg; Follow-up) | | 3,374 \| 52.26 | 2.76 (1.35) | .13-6.49 | 2.93 (1.34) | .13-6.49 | 2.50 (1.22) | .13-6.49 |
| IGF-1* (nmol/L; Baseline) | | 95 \| 1.47 | 2.77 (.34) | 1.10-4.19 | 2.78 (.34) | 1.10-4.19 | 2.78 (.32) | 1.61–3.85 |
| IGF-1* (nmol/L; Follow-up) | | 2,297 \| 35.58 | 2.78 (.27) | 1.61-4.06 | 2.78 (.27) | 1.61-4.06 | 2.79 (.30) | 1.61–4.06 |

Notes: ELSA, waves 4-6 (2008/09-2012/13); N = observations; % = percentage frequencies; SD = standard deviations; OSC = occupational social class; CRP = C-reactive protein; IGF-1 = Insulin-growth factor-1; * Log-transformed variable; I-N = immune and neuroendocrine.

***Table S2.*** **Correlations between immune and neuroendocrine biomarkers**

| | CRP | Fibrinogen | Cortisol | IGF-1 |
|---|---|---|---|---|
| CRP | 1<br>- | | | |
| Fibrinogen | 0.706*<br><0.001 | 1 | | |
| Cortisol | 0.273*<br><0.001 | 0.176*<br><0.001 | 1 | |
| IGF-1 | -0.163*<br><0.001 | -0.011<br>0.438 | 0.005<br>0.752 | 1 |

Notes: C-reactive protein (CRP); fibrinogen; insulin-growth factor-1 (IGF-1); * Significant at the 0.001 level

*Table S3.* **Seven Profile LPA model fit indices and predicted probability of profile membership (N = 4934)**

| **Criteria** | **One Profile** | **Two Profiles** | **Three Profiles** | **Four Profiles** | **Five Profiles** | **Six Profiles** | **Seven Profiles** |
|---|---|---|---|---|---|---|---|
| AIC | 36460.95 | 32478.36 | 30823.00 | 30574.08 | 30283.37 | 30214.23 | 30105.23 |
| AIC Difference (N) | - | 3982.59 | 1655.36 | 248.92 | 290.71 | 69.14 | 109.00 |
| AIC Difference (%) | - | 12.26 | 5.37 | 0.81 | 0.96 | 0.23 | 0.36 |
| BIC | 36512.98 | 32562.92 | 30940.07 | 30723.67 | 30465.48 | 30428.86 | 30352.38 |
| BIC Difference (N) | - | 3950.06 | 1622.85 | 216.40 | 258.19 | 36.62 | 76.48 |
| BIC Difference (%) | - | 12.13 | 5.25 | 0.70 | 0.85 | 0.12 | 0.25 |
| aBIC | 36487.56 | 32521.61 | 30882.87 | 30650.59 | 30376.51 | 30324.00 | 30231.63 |
| aBIC Difference (N) | - | 3965.95 | 1638.73 | 232.29 | 274.08 | 52.51 | 92.37 |
| aBIC Difference (%) | - | 12.19 | 5.31 | 0.76 | 0.90 | 0.17 | 0.31 |
| Entropy | - | 0.88 | 0.84 | 0.73 | 0.83 | 0.82 | 0.87 |
| Normalised Entropy | - | 0.84 | 0.78 | 0.64 | 0.77 | 0.77 | 0.83 |
| M Posterior Probabilities (SE) | - | 0.711 (.007) | 0.358 (.008) | 0.211 (.012) | 0.185 (.008) | 0.150 (.008) | 0.179 (.008) |
| | - | 0.289 (.007) | 0.399 (.008) | 0.218 (.011) | 0.225 (.008) | 0.185 (.010) | 0.042 (.009) |
| | - | - | 0.243 (.006) | 0.333 (.009) | 0.296 (.008) | 0.119 (.010) | 0.187 (.011) |
| | - | - | - | 0.238 (.006) | 0.073 (.006) | 0.258 (.010) | 0.296 (.008) |
| | - | - | - | - | 0.223 (.006) | 0.066 (.005) | 0.074 (.006) |
| | - | - | - | - | - | 0.221 (.006) | 0.003 (.001) |
| | - | - | - | - | - | - | 0.219 (.006) |
| N classes >5% | Yes | Yes | Yes | Yes | Yes | Yes | No |

Notes: AIC = Akaike information criterion; BIC = Bayesian information criterion; aBIC = adjusted Bayesian information criterion; N = number of observations; M = mean; SE = standard errors.

***Table S4.*** **Longitudinal associations of the stress score with immune and neuroendocrine biomarker profiles, with incremental model adjustment (N=4,934)**

| **Adjustments** | **Binary Stress Score** | | | | |
|---|---|---|---|---|---|
| | **RRR** | **SE** | **95% CI** | | ***p*** |
| ***Moderate-risk* Profile** | | | | | |
| Model 1: *Unadjusted* | 0.98 | 0.10 | 0.81 | 1.20 | 0.870 |
| Model 2: *Model 1 + baseline biomarkers* [a] | 1.01 | 0.11 | 0.83 | 1.24 | 0.898 |
| Model 3: *Model 2 + demographics & genetics* [b] | 1.14 | 0.12 | 0.93 | 1.41 | 0.213 |
| Model 3a: *Model 3 + socioeconomics* [b1] | 1.14 | 0.12 | 0.92 | 1.40 | 0.232 |
| Model 3b: *Model 3 + lifestyle* [b2] | 1.09 | 0.12 | 0.88 | 1.35 | 0.412 |
| Model 3c: *Model 3 + health* [b3] | 1.15 | 0.12 | 0.93 | 1.41 | 0.206 |
| Model 4: *Fully Adjusted* [c] | 1.10 | 0.12 | 0.89 | 1.35 | 0.401 |
| ***High-risk* Profile** | | | | | |
| Model 1: *Unadjusted* | 1.34 | 0.15 | 1.08 | 1.66 | 0.008 |
| Model 2: *Model 1 + baseline biomarkers* [a] | 1.42 | 0.18 | 1.10 | 1.83 | 0.007 |
| Model 3: *Model 2 + demographics & genetics* [b] | 1.80 | 0.24 | 1.39 | 2.35 | <0.001 |
| Model 3a: *Model 3 + socioeconomics* [b1] | 1.77 | 0.24 | 1.36 | 2.31 | <0.001 |
| Model 3b: *Model 3 + lifestyle* [b2] | 1.61 | 0.22 | 1.22 | 2.11 | 0.001 |
| Model 3c: *Model 3 + health* [b3] | 1.81 | 0.25 | 1.39 | 2.36 | <0.001 |
| Model 4: *Fully Adjusted* [c] | 1.61 | 0.22 | 1.23 | 2.12 | 0.001 |

Notes: The *low-risk* group is the reference; RRR = relative risk ratio; SE = standard errors; CI = confidence interval; *p* = significance value.

a Baseline biomarkers: C-reactive protein (CRP); fibrinogen; insulin-growth factor-1 (IGF-1).

b Demographic and genetic variables: age; sex; 10 principal components (PCs); CRP polygenic score (PGS); cortisol PGS; IGF-1PGS.

b1 Socioeconomics: education; occupational social status.

b2 Lifestyle: smoking status; alcohol consumption; physical activity.

b3 Health: chronic lung disease; coronary heart disease; abnormal heart rhythm; heart murmur; congestive heart failure; angina; hypertension; diabetes; cancer; Parkinson's; Alzheimer's; dementia; asthma; arthritis; osteoporosis; psychiatric disorder.

c Fully adjusted: CRP; fibrinogen; IGF-1; age; sex; 10 PCs; CRP PGS; cortisol PGS; IGF-1 PGS; education; occupational social status; smoking status; alcohol consumption; physical activity; health (i.e., chronic lung disease; coronary heart disease; abnormal heart rhythm; heart murmur; congestive heart failure; angina; hypertension; diabetes; cancer; Parkinson's; Alzheimer's; dementia; asthma; arthritis; osteoporosis; psychiatric disorder).

***Table S5.*** **Longitudinal associations of the stress score with immune and neuroendocrine biomarker profiles (N=4,934)**

| **Adjustments** | **Stress Score** | | | | |
|---|---|---|---|---|---|
| | **RRR** | **SE** | **95% CI** | | ***p*** |
| ***Moderate-risk* Profile** | | | | | |
| Model 1: *Unadjusted* | 1.08 | 0.05 | 1.00 | 1.18 | 0.766 |
| Model 2: *Model 1 + baseline biomarkers* [a] | 1.11 | 0.05 | 1.01 | 1.22 | 0.942 |
| Model 3: *Model 2 + demographics & genetics* [b] | 1.25 | 0.06 | 1.13 | 1.38 | 0.080 |
| Model 4: *Fully Adjusted* [c] | 1.19 | 0.06 | 1.07 | 1.31 | 0.175 |
| ***High-risk* Profile** | | | | | |
| Model 1: *Unadjusted* | 1.08 | 0.05 | 1.00 | 1.18 | 0.050 |
| Model 2: *Model 1 + baseline biomarkers* [a] | 1.11 | 0.05 | 1.01 | 1.22 | 0.031 |
| Model 3: *Model 2 + demographics & genetics* [b] | 1.25 | 0.06 | 1.13 | 1.38 | 0.000 |
| Model 4: *Fully Adjusted* [c] | 1.19 | 0.06 | 1.07 | 1.31 | 0.001 |

Notes: The *low-risk* group is the reference; RRR = relative risk ratio; SE = standard errors; CI = confidence interval; *p* = significance value.

a Baseline biomarkers: C-reactive protein (CRP); fibrinogen; insulin-growth factor-1 (IGF-1).

b Demographic and genetic variables: age; sex; 10 principal components (PCs); CRP polygenic score (PGS); cortisol PGS; IGF-1 PGS.

c All variables: CRP; fibrinogen; IGF-1; age; sex; 10 PCs; CRP PGS; cortisol PGS; IGF-1 PGS; education; occupational social status; smoking status; alcohol consumption; physical activity; health (i.e., chronic lung disease; coronary heart disease; abnormal heart rhythm; heart murmur; congestive heart failure; angina; hypertension; diabetes; cancer; Parkinson's; Alzheimer's; dementia; asthma; arthritis; osteoporosis; psychiatric disorder).

***Table S6a.*** **Longitudinal associations of financial strain with immune and neuroendocrine biomarker profiles (N=4,934)**

| Adjustments | Binary Financial Stress Score | | | | |
|---|---|---|---|---|---|
| | RRR | SE | 95% CI | | *p* |
| ***Moderate-risk* Profile** | | | | | |
| Model 1: *Unadjusted* | 1.28 | 0.12 | 1.07 | 1.53 | 0.007 |
| Model 2: *Model 1 + baseline biomarkers* [a] | 1.27 | 0.12 | 1.06 | 1.53 | 0.010 |
| Model 3: *Model 2 + demographics & genetics* [b] | 1.32 | 0.12 | 1.09 | 1.58 | 0.004 |
| Model 4: *Fully Adjusted* [c] | 1.23 | 0.12 | 1.02 | 1.48 | 0.033 |
| ***High-risk* Profile** | | | | | |
| Model 1: *Unadjusted* | 1.66 | 0.16 | 1.37 | 2.01 | <0.001 |
| Model 2: *Model 1 + baseline biomarkers* [a] | 1.66 | 0.19 | 1.33 | 2.09 | <0.001 |
| Model 3: *Model 2 + demographics & genetics* [b] | 1.79 | 0.21 | 1.42 | 2.26 | <0.001 |
| Model 4: *Fully Adjusted* [c] | 1.59 | 0.19 | 1.25 | 2.01 | <0.001 |

Notes: The *low-risk* group is the reference; RRR = relative risk ratio; SE = standard errors; CI = confidence interval; *p* = significance value.

a Baseline biomarkers: C-reactive protein (CRP); fibrinogen; insulin-growth factor-1 (IGF-1).

b Demographic and genetic variables: age; sex; 10 principal components (PCs); CRP polygenic score (PGS); cortisol PGS; IGF-1 PGS.

c All variables: CRP; fibrinogen; IGF-1; age; sex; 10 PCs; CRP PGS; cortisol PGS; IGF-1 PGS; education; occupational social status; smoking status; alcohol consumption; physical activity; health (i.e., chronic lung disease; coronary heart disease; abnormal heart rhythm; heart murmur; congestive heart failure; angina; hypertension; diabetes; cancer; Parkinson's; Alzheimer's; dementia; asthma; arthritis; osteoporosis; psychiatric disorder).

***Table S6b.*** **Longitudinal associations of care giving with immune and neuroendocrine biomarker profiles (N=4,934)**

| **Adjustments** | **Binary Care Giving Stress Score** | | | | |
|---|---|---|---|---|---|
| | **RRR** | **SE** | **95% CI** | | ***p*** |
| ***Moderate-risk* Profile** | | | | | |
| Model 1: *Unadjusted* | 1.05 | 0.14 | 0.82 | 1.36 | 0.685 |
| Model 2: *Model 1 + baseline biomarkers* [a] | 1.02 | 0.14 | 0.79 | 1.33 | 0.866 |
| Model 3: *Model 2 + demographics & genetics* [b] | 1.07 | 0.14 | 0.83 | 1.40 | 0.598 |
| Model 4: *Fully Adjusted* [c] | 1.10 | 0.15 | 0.84 | 1.43 | 0.484 |
| ***High-risk* Profile** | | | | | |
| Model 1: *Unadjusted* | 1.05 | 0.15 | 0.79 | 1.40 | 0.726 |
| Model 2: *Model 1 + baseline biomarkers* [a] | 1.02 | 0.17 | 0.73 | 1.42 | 0.930 |
| Model 3: *Model 2 + demographics & genetics* [b] | 1.17 | 0.21 | 0.83 | 1.65 | 0.371 |
| Model 4: *Fully Adjusted* [c] | 1.29 | 0.23 | 0.91 | 1.83 | 0.153 |

Notes: The *low-risk* group is the reference; RRR = relative risk ratio; SE = standard errors; CI = confidence interval; *p* = significance value.
a Baseline biomarkers: C-reactive protein (CRP); fibrinogen; insulin-growth factor-1 (IGF-1).
b Demographic and genetic variables: age; sex; 10 principal components (PCs); CRP polygenic score (PGS); cortisol PGS; IGF-1 PGS.
c All variables: CRP; fibrinogen; IGF-1; age; sex; 10 PCs; CRP PGS; cortisol PGS; IGF-1 PGS; education; occupational social status; smoking status; alcohol consumption; physical activity; health (i.e., chronic lung disease; coronary heart disease; abnormal heart rhythm; heart murmur; congestive heart failure; angina; hypertension; diabetes; cancer; Parkinson's; Alzheimer's; dementia; asthma; arthritis; osteoporosis; psychiatric disorder).

***Table S6c.*** **Longitudinal associations of disability with immune and neuroendocrine biomarker profiles (N=4,934)**

| **Adjustments** | **Binary Disability Stress Score** | | | | |
|---|---|---|---|---|---|
| | **RRR** | **SE** | **95% CI** | | ***p*** |
| ***Moderate-risk* Profile** | | | | | |
| Model 1: *Unadjusted* | 0.59 | 0.04 | 0.51 | 0.67 | <0.001 |
| Model 2: *Model 1 + baseline biomarkers* [a] | 0.65 | 0.04 | 0.57 | 0.75 | <0.001 |
| Model 3: *Model 2 + demographics & genetics* [b] | 0.74 | 0.05 | 0.64 | 0.85 | <0.001 |
| Model 4: *Fully Adjusted* [c] | 0.80 | 0.06 | 0.69 | 0.92 | 0.002 |
| ***High-risk* Profile** | | | | | |
| Model 1: *Unadjusted* | 0.33 | 0.03 | 0.28 | 0.39 | <0.001 |
| Model 2: *Model 1 + baseline biomarkers* [a] | 0.46 | 0.04 | 0.39 | 0.55 | <0.001 |
| Model 3: *Model 2 + demographics & genetics* [b] | 0.55 | 0.05 | 0.45 | 0.66 | <0.001 |
| Model 4: *Fully Adjusted* [c] | 0.70 | 0.07 | 0.58 | 0.86 | <0.001 |

Notes: The *low-risk* group is the reference; RRR = relative risk ratio; SE = standard errors; CI = confidence interval; *p* = significance value.
a Baseline biomarkers: C-reactive protein (CRP); fibrinogen; insulin-growth factor-1 (IGF-1).
b Demographic and genetic variables: age; sex; 10 principal components (PCs); CRP polygenic score (PGS); cortisol PGS; IGF-1 PGS.
c All variables: CRP; fibrinogen; IGF-1; age; sex; 10 PCs; CRP PGS; cortisol PGS; IGF-1 PGS; education; occupational social status; smoking status; alcohol consumption; physical activity; health (i.e., chronic lung disease; coronary heart disease; abnormal heart rhythm; heart murmur; congestive heart failure; angina; hypertension; diabetes; cancer; Parkinson's; Alzheimer's; dementia; asthma; arthritis; osteoporosis; psychiatric disorder).

***Table S6d.*** **Longitudinal associations of limiting longstanding illness with immune and neuroendocrine biomarker profiles (N=4,934)**

| **Adjustments** | **Binary Limiting Longstanding Illness Stress Score** | | | | |
|---|---|---|---|---|---|
| | **RRR** | **SE** | **95% CI** | | ***p*** |
| ***Moderate-risk* Profile** | | | | | |
| Model 1: *Unadjusted* | 1.46 | 0.11 | 1.26 | 1.69 | <0.001 |
| Model 2: *Model 1 + baseline biomarkers* [a] | 1.34 | 0.10 | 1.15 | 1.55 | <0.001 |
| Model 3: *Model 2 + demographics & genetics* [b] | 1.25 | 0.10 | 1.07 | 1.46 | 0.004 |
| Model 4: *Fully Adjusted* [c] | 1.14 | 0.09 | 0.97 | 1.34 | 0.112 |
| ***High-risk* Profile** | | | | | |
| Model 1: *Unadjusted* | 2.64 | 0.21 | 2.26 | 3.10 | <0.001 |
| Model 2: *Model 1 + baseline biomarkers* [a] | 2.01 | 0.19 | 1.67 | 2.42 | <0.001 |
| Model 3: *Model 2 + demographics & genetics* [b] | 1.81 | 0.18 | 1.50 | 2.18 | <0.001 |
| Model 4: *Fully Adjusted* [c] | 1.34 | 0.14 | 1.10 | 1.65 | 0.005 |

Notes: The *low-risk* group is the reference; RRR = relative risk ratio; SE = standard errors; CI = confidence interval; *p* = significance value.
a Baseline biomarkers: C-reactive protein (CRP); fibrinogen; insulin-growth factor-1 (IGF-1).
b Demographic and genetic variables: age; sex; 10 principal components (PCs); CRP polygenic score (PGS); cortisol PGS; IGF-1 PGS.
c All variables: CRP; fibrinogen; IGF-1; age; sex; 10 PCs; CRP PGS; cortisol PGS; IGF-1 PGS; education; occupational social status; smoking status; alcohol consumption; physical activity; health (i.e., chronic lung disease; coronary heart disease; abnormal heart rhythm; heart murmur; congestive heart failure; angina; hypertension; diabetes; cancer; Parkinson's; Alzheimer's; dementia; asthma; arthritis; osteoporosis; psychiatric disorder).

***Table S6e.*** **Longitudinal associations of bereavement with immune and neuroendocrine biomarker profiles (N=4,934)**

| **Adjustments** | **Binary Bereavement Stress Score** | | | | |
|---|---|---|---|---|---|
| | **RRR** | **SE** | **95% CI** | | ***p*** |
| ***Moderate-risk* Profile** | | | | | |
| Model 1: *Unadjusted* | 1.06 | 0.07 | 0.93 | 1.21 | 0.397 |
| Model 2: *Model 1 + baseline biomarkers* [a] | 1.07 | 0.07 | 0.93 | 1.22 | 0.354 |
| Model 3: *Model 2 + demographics & genetics* [b] | 1.16 | 0.08 | 1.01 | 1.33 | 0.040 |
| Model 4: *Fully Adjusted* [c] | 1.18 | 0.08 | 1.02 | 1.36 | 0.022 |
| ***High-risk* Profile** | | | | | |
| Model 1: *Unadjusted* | 1.11 | 0.08 | 0.96 | 1.29 | 0.178 |
| Model 2: *Model 1 + baseline biomarkers* [a] | 1.10 | 0.10 | 0.93 | 1.31 | 0.273 |
| Model 3: *Model 2 + demographics & genetics* [b] | 1.25 | 0.12 | 1.04 | 1.50 | 0.016 |
| Model 4: *Fully Adjusted* [c] | 1.26 | 0.12 | 1.04 | 1.52 | 0.016 |

Notes: The *low-risk* group is the reference; RRR = relative risk ratio; SE = standard errors; CI = confidence interval; *p* = significance value.

a Baseline biomarkers: C-reactive protein (CRP); fibrinogen; insulin-growth factor-1 (IGF-1).

b Demographic and genetic variables: age; sex; 10 principal components (PCs); CRP polygenic score (PGS); cortisol PGS; IGF-1 PGS.

c All variables: CRP; fibrinogen; IGF-1; age; sex; 10 PCs; CRP PGS; cortisol PGS; IGF-1 PGS; education; occupational social status; smoking status; alcohol consumption; physical activity; health (i.e., chronic lung disease; coronary heart disease; abnormal heart rhythm; heart murmur; congestive heart failure; angina; hypertension; diabetes; cancer; Parkinson's; Alzheimer's; dementia; asthma; arthritis; osteoporosis; psychiatric disorder).

***Table S6f.*** **Longitudinal associations of divorce with immune and neuroendocrine biomarker profiles (N=4,934)**

| Adjustments | Binary Divorce Stress Score | | | | |
|---|---|---|---|---|---|
| | **RRR** | **SE** | **95% CI** | | ***p*** |
| ***Moderate-risk* Profile** | | | | | |
| Model 1: *Unadjusted* | 1.03 | 0.12 | 0.82 | 1.29 | 0.810 |
| Model 2: *Model 1 + baseline biomarkers* [a] | 1.05 | 0.13 | 0.83 | 1.32 | 0.698 |
| Model 3: *Model 2 + demographics & genetics* [b] | 1.10 | 0.13 | 0.87 | 1.39 | 0.441 |
| Model 4: *Fully Adjusted* [c] | 0.98 | 0.12 | 0.77 | 1.25 | 0.886 |
| ***High-risk* Profile** | | | | | |
| Model 1: *Unadjusted* | 1.27 | 0.16 | 0.99 | 1.62 | 0.060 |
| Model 2: *Model 1 + baseline biomarkers* [a] | 1.31 | 0.20 | 0.98 | 1.76 | 0.069 |
| Model 3: *Model 2 + demographics & genetics* [b] | 1.52 | 0.23 | 1.13 | 2.05 | 0.006 |
| Model 4: *Fully Adjusted* [c] | 1.20 | 0.19 | 0.88 | 1.64 | 0.243 |

Notes: The *low-risk* group is the reference; RRR = relative risk ratio; SE = standard errors; CI = confidence interval; *p* = significance value.

a Baseline biomarkers: C-reactive protein (CRP); fibrinogen; insulin-growth factor-1 (IGF-1).

b Demographic and genetic variables: age; sex; 10 principal components (PCs); CRP polygenic score (PGS); cortisol PGS; IGF-1 PGS.

c All variables: CRP; fibrinogen; IGF-1; age; sex; 10 PCs; CRP PGS; cortisol PGS; IGF-1 PGS; education; occupational social status; smoking status; alcohol consumption; physical activity; health (i.e., chronic lung disease; coronary heart disease; abnormal heart rhythm; heart murmur; congestive heart failure; angina; hypertension; diabetes; cancer; Parkinson's; Alzheimer's; dementia; asthma; arthritis; osteoporosis; psychiatric disorder).

*Table S7.* **Longitudinal associations of stress with immune and neuroendocrine biomarker profiles, excluding measures for disability and limiting longstanding illness (N=4,934)**

| **Adjustments** | **Reduced Binary Stress Score** | | | | |
|---|---|---|---|---|---|
| | **RRR** | **SE** | **95% CI** | | ***p*** |
| ***Moderate-risk* Profile** | | | | | |
| Model 1: *Unadjusted* | 1.04 | 0.10 | 0.86 | 1.26 | 0.698 |
| Model 2: *Model 1 + baseline biomarkers* [a] | 1.05 | 0.11 | 0.86 | 1.28 | 0.646 |
| Model 3: *Model 2 + demographics & genetics* [b] | 1.17 | 0.12 | 0.95 | 1.43 | 0.141 |
| Model 4: *Fully Adjusted* [c] | 1.11 | 0.12 | 0.90 | 1.36 | 0.335 |
| ***High-risk* Profile** | | | | | |
| Model 1: *Unadjusted* | 1.47 | 0.15 | 1.19 | 1.80 | <0.001 |
| Model 2: *Model 1 + baseline biomarkers* [a] | 1.52 | 0.19 | 1.19 | 1.94 | 0.001 |
| Model 3: *Model 2 + demographics & genetics* [b] | 1.89 | 0.24 | 1.47 | 2.43 | <0.001 |
| Model 4: *Fully Adjusted* [c] | 1.71 | 0.23 | 1.32 | 2.22 | <0.001 |

| **Adjustments** | **Reduced Stress Score** | | | | |
|---|---|---|---|---|---|
| | **RRR** | **SE** | **95% CI** | | ***p*** |
| ***Moderate-risk* Profile** | | | | | |
| Model 1: *Unadjusted* | 1.10 | 0.05 | 1.01 | 1.21 | 0.029 |
| Model 2: *Model 1 + baseline biomarkers* [a] | 1.11 | 0.05 | 1.01 | 1.21 | 0.033 |
| Model 3: *Model 2 + demographics & genetics* [b] | 1.18 | 0.06 | 1.07 | 1.30 | 0.001 |
| Model 4: *Fully Adjusted* [c] | 1.15 | 0.06 | 1.04 | 1.27 | 0.004 |
| ***High-risk* Profile** | | | | | |
| Model 1: *Unadjusted* | 1.25 | 0.06 | 1.13 | 1.38 | <0.001 |
| Model 2: *Model 1 + baseline biomarkers* [a] | 1.25 | 0.08 | 1.11 | 1.41 | <0.001 |
| Model 3: *Model 2 + demographics & genetics* [b] | 1.42 | 0.09 | 1.26 | 1.61 | <0.001 |
| Model 4: *Fully Adjusted* [c] | 1.35 | 0.09 | 1.19 | 1.53 | <0.001 |

Notes: The *low-risk* group is the reference; RRR = relative risk ratio; SE = standard errors; CI = confidence interval; $p$ = significance value.

a Baseline biomarkers: C-reactive protein (CRP); fibrinogen; insulin-growth factor-1 (IGF-1).

b Demographic and genetic variables: age; sex; 10 principal components (PCs); CRP polygenic score (PGS); cortisol PGS; IGF-1 PGS.

c All variables: CRP; fibrinogen; IGF-1; age; sex; 10 PCs; CRP PGS; cortisol PGS; IGF-1 PGS; education; occupational social status; smoking status; alcohol consumption; physical activity; health (i.e., chronic lung disease; coronary heart disease; abnormal heart rhythm; heart murmur; congestive heart failure; angina; hypertension; diabetes; cancer; Parkinson's; Alzheimer's; dementia; asthma; arthritis; osteoporosis; psychiatric disorder).

*Table S8a.* **Longitudinal associations of stress with immune and neuroendocrine biomarker profiles stratified by median age (65 years)**

| Adjustments | Binary Stress Score \| < Mdn Age (N=2,437) | | | | | Binary Stress Score \| ≥ Mdn Age (N=2,497) | | | | |
|---|---|---|---|---|---|---|---|---|---|---|
| | RRR | SE | 95% CI | | *p* | RRR | SE | 95% CI | | *p* |
| ***Moderate-risk* Profile** | | | | | | | | | | |
| Model 1: *Unadjusted* | 1.06 | 0.13 | 0.83 | 1.35 | 0.660 | 1.10 | 0.21 | 0.76 | 1.59 | 0.624 |
| Model 2: *Model 1 + baseline biomarkers* [a] | 1.13 | 0.15 | 0.88 | 1.45 | 0.350 | 1.08 | 0.21 | 0.74 | 1.58 | 0.687 |
| Model 3: *Model 2 + demographics & genetics* [b] | 1.13 | 0.15 | 0.88 | 1.46 | 0.343 | 1.16 | 0.23 | 0.79 | 1.70 | 0.458 |
| Model 4: *Fully Adjusted* [c] | 1.08 | 0.14 | 0.83 | 1.39 | 0.583 | 1.12 | 0.22 | 0.76 | 1.65 | 0.581 |
| ***High-risk* Profile** | | | | | | | | | | |
| Model 1: *Unadjusted* | 1.26 | 0.18 | 0.95 | 1.67 | 0.117 | 2.05 | 0.38 | 1.43 | 2.95 | <0.001 |
| Model 2: *Model 1 + baseline biomarkers* [a] | 1.40 | 0.24 | 0.99 | 1.96 | 0.056 | 1.98 | 0.42 | 1.31 | 3.00 | 0.001 |
| Model 3: *Model 2 + demographics & genetics* [b] | 1.37 | 0.24 | 0.96 | 1.93 | 0.079 | 2.29 | 0.51 | 1.49 | 3.53 | <0.001 |
| Model 4: *Fully Adjusted* [c] | 1.18 | 0.22 | 0.83 | 1.69 | 0.354 | 2.13 | 0.49 | 1.36 | 3.34 | 0.001 |

Notes: The *low-risk* group is the reference; < = less than; ≥ = greater than or equal to; Mdn = Median; RRR = relative risk ratio; SE = standard errors; CI = confidence interval; *p* = significance value.

a Baseline biomarkers: C-reactive protein (CRP); fibrinogen; insulin-growth factor-1 (IGF-1).

b Demographic and genetic variables: age; sex; 10 principal components (PCs); CRP polygenic score (PGS); cortisol PGS; IGF-1 PGS.

c All variables: CRP; fibrinogen; IGF-1; age; sex; 10 PCs; CRP PGS; cortisol PGS; IGF-1 PGS; education; occupational social status; smoking status; alcohol consumption; physical activity; health (i.e., chronic lung disease; coronary heart disease; abnormal heart rhythm; heart murmur; congestive heart failure; angina; hypertension; diabetes; cancer; Parkinson's; Alzheimer's; dementia; asthma; arthritis; osteoporosis; psychiatric disorder).

***Table S8b.*** **The moderated effective of age on longitudinal associations between stress and immune and neuroendocrine biomarker profiles (N=4,934)**

| **Adjustments** | **Stress Score \| Age** | | | | |
|---|---|---|---|---|---|
| | **RRR** | **SE** | **95% CI** | | ***p*** |
| ***Moderate-risk* Profile** | | | | | |
| Model 1: *Unadjusted* | 1.00 | 0.00 | 0.99 | 1.01 | 0.445 |
| Model 2: *Model 1 + baseline biomarkers* [a] | 1.00 | 0.00 | 0.99 | 1.00 | 0.229 |
| Model 3: *Model 2 + demographics & genetics* [b] | 0.99 | 0.01 | 0.99 | 1.00 | 0.202 |
| Model 4: *Fully Adjusted* [c] | 0.99 | 0.01 | 0.99 | 1.00 | 0.172 |
| ***High-risk* Profile** | | | | | |
| Model 1: *Unadjusted* | 1.01 | 0.01 | 1.00 | 1.02 | 0.205 |
| Model 2: *Model 1 + baseline biomarkers* [a] | 1.00 | 0.01 | 0.99 | 1.01 | 0.829 |
| Model 3: *Model 2 + demographics & genetics* [b] | 1.00 | 0.01 | 0.99 | 1.01 | 0.801 |
| Model 4: *Fully Adjusted* [c] | 1.00 | 0.01 | 0.99 | 1.01 | 0.913 |

*Table S9a.* **Longitudinal associations of stress with immune and neuroendocrine biomarker profiles stratified by sex**

| **Adjustments** | **Binary Stress Score \| Male (N=2,235)** | | | | | **Binary Stress Score \| Female (N=2,699)** | | | | |
|---|---|---|---|---|---|---|---|---|---|---|
| | **RRR** | **SE** | **95% CI** | | ***p*** | **RRR** | **SE** | **95% CI** | | ***p*** |
| ***Moderate-risk* Profile** | | | | | | | | | | |
| Model 1: *Unadjusted* | 0.90 | 0.15 | 0.66 | 1.25 | 0.540 | 1.01 | 0.13 | 0.78 | 1.31 | 0.922 |
| Model 2: *Model 1 + baseline biomarkers* [a] | 0.96 | 0.16 | 0.69 | 1.33 | 0.790 | 1.03 | 0.14 | 0.79 | 1.35 | 0.818 |
| Model 3: *Model 2 + demographics & genetics* [b] | 1.09 | 0.19 | 0.78 | 1.53 | 0.611 | 1.16 | 0.16 | 0.89 | 1.52 | 0.280 |
| Model 4: *Fully Adjusted* [c] | 0.97 | 0.17 | 0.69 | 1.37 | 0.857 | 1.16 | 0.16 | 0.88 | 1.53 | 0.289 |
| ***High-risk* Profile** | | | | | | | | | | |
| Model 1: *Unadjusted* | 1.32 | 0.23 | 0.94 | 1.84 | 0.112 | 1.34 | 0.19 | 1.01 | 1.76 | 0.042 |
| Model 2: *Model 1 + baseline biomarkers* [a] | 1.46 | 0.30 | 0.97 | 2.18 | 0.069 | 1.41 | 0.24 | 1.02 | 1.97 | 0.040 |
| Model 3: *Model 2 + demographics & genetics* [b] | 1.82 | 0.39 | 1.19 | 2.77 | 0.006 | 1.78 | 0.31 | 1.26 | 2.50 | 0.001 |
| Model 4: *Fully Adjusted* [c] | 1.48 | 0.33 | 0.95 | 2.29 | 0.080 | 1.66 | 0.30 | 1.17 | 2.37 | 0.005 |

Notes: The *low-risk* group is the reference; RRR = relative risk ratio; SE = standard errors; CI = confidence interval; *p* = significance value.

a Baseline biomarkers: C-reactive protein (CRP); fibrinogen; insulin-growth factor-1 (IGF-1).

b Demographic and genetic variables: age; sex; 10 principal components (PCs); CRP polygenic score (PGS); cortisol PGS; IGF-1 PGS.

c All variables: CRP; fibrinogen; IGF-1; age; sex; 10 PCs; CRP PGS; cortisol PGS; IGF-1 PGS; education; occupational social status; smoking status; alcohol consumption; physical activity; health (i.e., chronic lung disease; coronary heart disease; abnormal heart rhythm; heart murmur; congestive heart failure; angina; hypertension; diabetes; cancer; Parkinson's; Alzheimer's; dementia; asthma; arthritis; osteoporosis; psychiatric disorder).

***Table S9b.*** **The moderated effective of sex on longitudinal associations between stress and immune and neuroendocrine biomarker profiles (N=4,934)**

| Adjustments | Stress Score \| Sex | | | | |
|---|---|---|---|---|---|
| | **RRR** | **SE** | **95% CI** | | ***p*** |
| ***Moderate-risk* Profile** | | | | | |
| Model 1: *Unadjusted* | 1.03 | 0.08 | 0.89 | 1.19 | 0.704 |
| Model 2: *Model 1 + baseline biomarkers* [a] | 1.05 | 0.08 | 0.91 | 1.23 | 0.495 |
| Model 3: *Model 2 + demographics & genetics* [b] | 1.11 | 0.09 | 0.95 | 1.29 | 0.189 |
| Model 4: *Fully Adjusted* [c] | 1.09 | 0.09 | 0.93 | 1.27 | 0.299 |
| ***High-risk* Profile** | | | | | |
| Model 1: *Unadjusted* | 1.03 | 0.09 | 0.87 | 1.21 | 0.742 |
| Model 2: *Model 1 + baseline biomarkers* [a] | 1.08 | 0.11 | 0.89 | 1.31 | 0.469 |
| Model 3: *Model 2 + demographics & genetics* [b] | 1.15 | 0.12 | 0.94 | 1.41 | 0.163 |
| Model 4: *Fully Adjusted* [c] | 1.13 | 0.12 | 0.92 | 1.39 | 0.239 |

***Table S10.*** **Longitudinal associations of stress with immune and neuroendocrine biomarker profiles, with complete case data (N=1,677)**

| Adjustments | Binary Stress Score | | | | |
|---|---|---|---|---|---|
| | **RRR** | **SE** | **95% CI** | | ***p*** |
| ***Moderate-risk* Profile** | | | | | |
| Model 1: *Unadjusted* | 1.23 | 0.20 | 0.89 | 1.70 | 0.210 |
| Model 2: *Model 1 + baseline biomarkers* [a] | 1.33 | 0.25 | 0.93 | 1.91 | 0.117 |
| Model 3: *Model 2 + demographics & genetics* [b] | 1.38 | 0.26 | 0.96 | 1.99 | 0.084 |
| Model 4: *Fully Adjusted* [c] | 1.33 | 0.25 | 0.91 | 1.93 | 0.136 |
| ***High-risk* Profile** | | | | | |
| Model 1: *Unadjusted* | 1.22 | 0.34 | 0.71 | 2.11 | 0.473 |
| Model 2: *Model 1 + baseline biomarkers* [a] | 1.35 | 0.41 | 0.74 | 2.46 | 0.330 |
| Model 3: *Model 2 + demographics & genetics* [b] | 1.44 | 0.45 | 0.78 | 2.65 | 0.239 |
| Model 4: *Fully Adjusted* [c] | 1.23 | 0.40 | 0.65 | 2.32 | 0.524 |

| Adjustments | Stress Score | | | | |
|---|---|---|---|---|---|
| | **RRR** | **SE** | **95% CI** | | ***p*** |
| ***Moderate-risk* Profile** | | | | | |
| Model 1: *Unadjusted* | 1.09 | 0.07 | 0.97 | 1.23 | 0.141 |
| Model 2: *Model 1 + baseline biomarkers* [a] | 1.11 | 0.07 | 0.97 | 1.26 | 0.125 |
| Model 3: *Model 2 + demographics & genetics* [b] | 1.13 | 0.08 | 0.99 | 1.29 | 0.071 |
| Model 4: *Fully Adjusted* [c] | 1.12 | 0.08 | 0.98 | 1.29 | 0.096 |
| ***High-risk* Profile** | | | | | |
| Model 1: *Unadjusted* | 1.13 | 0.12 | 0.92 | 1.38 | 0.236 |
| Model 2: *Model 1 + baseline biomarkers* [a] | 1.14 | 0.13 | 0.92 | 1.43 | 0.241 |
| Model 3: *Model 2 + demographics & genetics* [b] | 1.19 | 0.14 | 0.95 | 1.49 | 0.141 |
| Model 4: *Fully Adjusted* [c] | 1.09 | 0.13 | 0.86 | 1.38 | 0.492 |

Notes: The *low-risk* group is the reference; RRR = relative risk ratio; SE = standard errors; CI = confidence interval; *p* = significance value.

a Baseline biomarkers: C-reactive protein (CRP); fibrinogen; insulin-growth factor-1 (IGF-1).

b Demographic and genetic variables: age; sex; 10 principal components (PCs); CRP polygenic score (PGS); cortisol PGS; IGF-1 PGS.

c All variables: CRP; fibrinogen; IGF-1; age; sex; 10 PCs; CRP PGS; cortisol PGS; IGF-1 PGS; education; occupational social status; smoking status; alcohol consumption; physical activity; health (i.e., chronic lung disease; coronary heart disease; abnormal heart rhythm; heart murmur; congestive heart failure; angina; hypertension; diabetes; cancer; Parkinson's; Alzheimer's; dementia; asthma; arthritis; osteoporosis; psychiatric disorder).
